# Baseline Gut Microbiome-Metabolome Signatures Are Associated with Drinking Severity and Reduction Following Dutasteride Treatment in Alcohol Use Disorder

**DOI:** 10.64898/2026.05.26.26354041

**Authors:** L.R. Dedon, D.J. Lee, Q. Lin, H. Yuan, J. Chi, L. Li, H. Gu, H. Tennen, J.M. Covault, Y. Zhou

**Author notes:** Authors contributed equally (L.R. Dedon, D.J. Lee).

## Abstract

The gut microbiome has been implicated in alcohol use disorder (AUD), but its relationship to drinking intensity and treatment response remains poorly understood. We conducted a longitudinal multi-omics analysis of stool samples collected at baseline and endpoint (after 12 weeks) from 122 participants enrolled in a double-blind, placebo-controlled trial of dutasteride for AUD. Gut microbiome composition was characterized using 16S rRNA gene sequencing, and fecal metabolites were measured by LC-MS–based metabolomics. At baseline, drinking intensity was associated with increasingly lower microbial richness. Genera in the class *Clostridia* emerged as key microbial hubs associated with drinking intensity in an age- and sex-dependent manner. Drinking intensity promoted co-enrichment of [*Ruminococcus*] *gnavus* group and [*Clostridium*] *inocuum* group with amino acid catabolites, as well as the co-depletion of diverse *Clostridia* taxa and lipid metabolites. Dutasteride treatment and drinking reduction had minimal impact on gut microbiome composition. Random forest models integrating baseline clinical, microbiome, and metabolome data improved the classification of clinically meaningful drinking reduction compared to models using clinical data alone. These findings show that a coupled baseline gut microbiome-metabolome signature is associated with drinking intensity and future treatment response in AUD, highlighting the potential for multi-omics integration to inform precision treatment approaches.

## Background

Alcohol use disorder (AUD) persists as an unresolved public health challenge, affecting millions of individuals across age groups and sexes.^2, 3^ AUD is a risk factor for developing many adverse health conditions, including several cancers, cognitive decline, gastrointestinal disorders, and liver disease. ^4–7^ A growing body of evidence suggests that the gut microbiome is implicated in AUD.^8–11^ Long-term drinking is associated with distinct changes in gut microbiome composition.^12–16^ However, most existing studies have focused on relatively small case-control studies comparing the microbiomes of healthy controls and individuals with AUD, whereas the relationship between the microbiome and drinking intensity within AUD populations is less explored. In addition, AUD phenotype and gut microbiome composition and function are strongly shaped by demographic factors such as age, sex, and psychological symptoms.^17–25^ Despite this, these important parameters are not adequately accounted for in many microbiome analyses of AUD cohorts, which can potentially introduce bias in the identification of microbiota associated with drinking behavior and contribute to inconsistent findings across studies.

Beyond microbial composition, the gut metabolome reflects the functional state of host–microbiome interactions and may offer additional insight into health effects associated with alcohol.^26–28^ Gut metabolites capture the integrated metabolic activity of the microbiome, host physiology, and dietary inputs, enabling characterization of phenotypic variation that may not be fully resolved by taxonomic profiling alone. Given the close relationship between microbial communities and their metabolic output, alcohol-related changes in the gut microbiome are likely accompanied by coordinated shifts in the gut metabolome. Prior work has linked AUD to alterations in fecal short-chain fatty acid production.^9, 13, 29^ Circulating tryptophan metabolites, which are strongly influenced by microbial metabolism, have also been correlated with alcohol-associated dysbiosis and co-occurring psychiatric conditions.^30–34^ However, the effects of drinking behavior and AUD intensity on the gut metabolome have yet to be comprehensively defined. Integrated analysis of the gut microbiome and metabolome is therefore an important area for inquiry that may help clarify the metabolic changes underlying alcohol-related gut dysfunction.

Treatment outcomes for pharmacological and behavioral treatment of AUD are highly variable across individuals. Despite this, predictive models for treatment response based on demographic and drinking clinical data have demonstrated modest accuracy, with sex-specific differences.^35, 36^ To further elucidate what mechanisms drive individual treatment success, there is a critical need to identify additional biological factors that influence both initial drinking behavior and treatment response. The gut microbiome is a promising candidate for such work due to its reciprocal effects on host metabolism and immune regulation.^37, 38^ Baseline microbiome composition has been shown to influence treatment response to chemotherapy, nutritional intervention, and vaccination.^39–42^ Likewise, baseline -omics profiles among patients with AUD, as well as changes during treatment, may contain relevant biological markers for treatment outcomes and could provide a foundation to establish more effective, personalized strategies for AUD intervention.

We hypothesized that baseline gut microbiome and metabolome features would be associated with baseline drinking intensity and would predict changes in drinking among patients undergoing treatment for AUD. We tested these hypotheses by conducting a secondary multi-omics analysis of stool samples from a large randomized clinical trial in which 12 weeks of dutasteride treatment significantly reduced drinking and heavy drinking in patients with AUD.^1^ We find that genera in the class *Clostridia* act as microbial “hub taxa” that are strongly associated with baseline drinking intensity, with effects varying by sex and age: most notably, older patients exhibit smaller effects overall. Microbiome–metabolome co-associations show that higher drinking intensity is associated with increases in a subset of taxa involved in tryptophan and amino acid metabolism and reductions in a larger group associated with lipid metabolism. Furthermore, integrating baseline microbiome and metabolome data into a clinical random forest model improves its ability to classify clinically meaningful drinking reduction. Our findings suggest that drinking intensity shapes coordinated shifts in the gut microbiome and metabolome within AUD cohorts, and that baseline multi-omics data may serve as relevant predictors of treatment response.

## Results

### Study Participants

Participants with AUD received 12 weeks of dutasteride or placebo treatment to reduce drinking (ClinicalTrials.gov identifier: NCT04098302)^1^. The secondary analysis described here included 122 out of 155 participants who provided baseline stool samples (Fig 1a). Of these, 77 participants also provided endpoint stool samples.^1^ The demographic and clinical characteristics of the 122 participants at baseline are summarized in Table 1. Thirty-eight percent of participants were female. Participant age ranged from 37 to 70 years. Nearly all participants (94%) identified as white/non-Hispanic. The only significant difference between the dutasteride and placebo groups was that a larger fraction of participants reported the use of prescribed psychotropic medication in the dutasteride group as compared to placebo (46% and 23%, respectively, p = 0.013).

**Figure 1:**
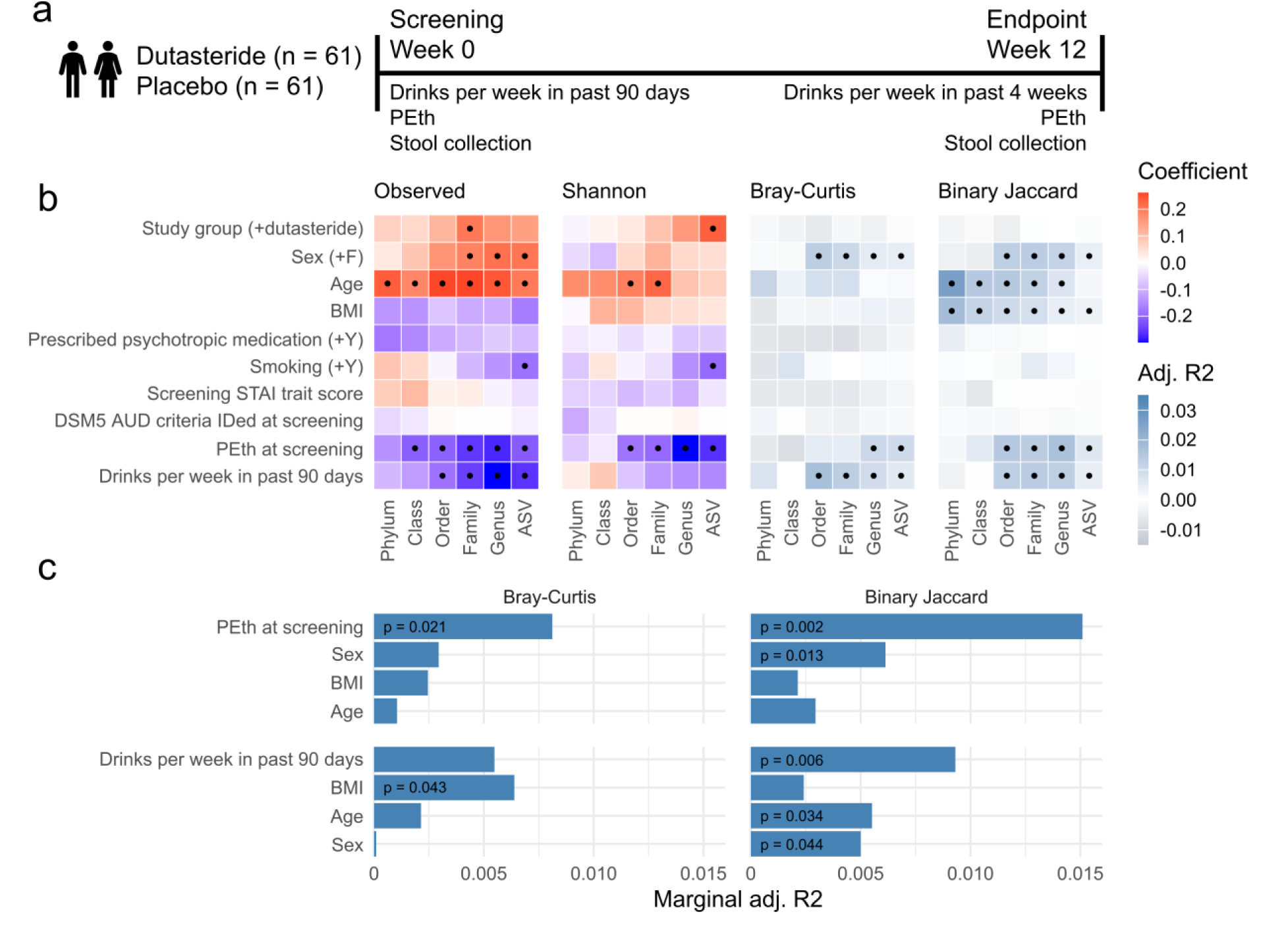
Study design and microbiome diversity associations with clinical characteristics. (a) Simplified schematic of study design and drinking metadata and biological sample collection. (b) Heatmap showing associations of alpha (richness, Shannon diversity) and beta (Bray-Curtis and binary Jaccard) diversity with single-variable clinical characteristics at each taxonomic level (x-axis). Dots indicate statistically significant associations (PERMANOVA p < 0.05). (c) Bar plot of PERMANOVA (controlled for covariates) R² values at genus level for clinical variables significantly associated with single-variable beta diversity.

**Table 1:** Demographic and clinical factors for participants in this study.

| Variable | Overall<br>N = 122<sup>1</sup> | Placebo<br>N = 61<sup>1</sup> | Dutasteride<br>N = 61<sup>1</sup> | p-value |
| --- | --- | --- | --- | --- |
| Sex, n, (%) |  |  |  | 0.4<sup>2</sup> |
| Male | 76, (62%) | 41, (67%) | 35, (57%) |  |
| Female | 46, (38%) | 20, (33%) | 26, (43%) |  |
| Age, Mean, (SD) | 57, (9) | 56, (9) | 57, (9) | 0.5<sup>3</sup> |
| BMI, Mean, (SD) | 29.3, (5.8) | 29.1, (4.6) | 29.5, (6.9) | 0.7<sup>3</sup> |
| Prescribed psychotropic medication, n, (%) | 42, (34%) | 14, (23%) | 28, (46%) | 0.013<sup>2</sup> |
| Smoking, n, (%) | 15, (12%) | 9, (15%) | 6, (9.8%) | 0.6<sup>2</sup> |
| Primary race, n, (%) |  |  |  | 0.11<sup>2</sup> |
| White | 115, (94%) | 60, (98%) | 55, (90%) |  |
| Black | 7, (5.7%) | 1, (1.6%) | 6, (9.8%) |  |
| Screening STAI trait score, Mean, (SD) | 39, (12) | 37, (11) | 41, (12) | 0.077<sup>3</sup> |
| DSM5 AUD criteria identified at screening, Mean, (SD) | 5.75, (2.29) | 5.66, (2.17) | 5.84, (2.42) | 0.7<sup>3</sup> |
| PEth at screening, Mean, (SD) | 353, (290) | 372, (326) | 335, (254) | 0.5<sup>3</sup> |
| Average drinks per week at screening, Mean, (SD) | 45, (22) | 45, (22) | 44, (23) | 0.8<sup>3</sup> |

<sup>1</sup>Frequency, (%) or Mean, (SD)
<sup>2</sup>Fisher's exact test
<sup>3</sup>Welch Two Sample t-test

Blood phosphatidylethanol (PEth) and the self-reported average drinks per week in the 90 days preceding the screening visit did not significantly vary between treatment groups (p = 0.5 and 0.8, respectively). PEth is a unique phospholipid formed only when alcohol is present in the bloodstream and serves as a highly specific biomarker of alcohol consumption in the past 2 to 3 weeks.^43^ We will henceforth refer to the self-reported average drinks per week in the 90 days preceding the screening visit as baseline drinks per week. Using the same generalized linear mixed model (GLMM) approach from the original clinical trial,^1^ we confirmed that the subgroup of 122 participants in this study experienced a significant reduction in drinking (p = 0.04) among dutasteride-treated participants compared to the placebo group.

### The Baseline Gut Microbiome Community Is Influenced by Sex, Age, and Drinking Intensity

A total of 5,242,663 sequence reads passing QC were obtained from the 122 participants, with an average of 42,973 ± 22,745 reads per sample. From these sequences, taxa from across 7 phyla, 22 orders, 36 families, 105 genera, and 167 ASVs were included in the analysis. Bar plots of the relative abundance of the microbiome for all participants stratified by treatment are given in Supporting Fig. 1.

We first examined whether any individual demographic or clinical factors were associated with the overall baseline microbiome community as measured by alpha and beta diversity. Alpha diversity was positively correlated with age, as seen in observed features (richness) at all taxonomic levels and in Shannon diversity at the order and family levels (genus richness p = 0.007, Shannon diversity p = 0.011 at family level, Fig. 1b). Female participants exhibited higher richness than males at the family level and below (p = 0.019; p-values are reported at the genus level unless mentioned otherwise). Binary Jaccard distances significantly varied with age and BMI across all taxonomic levels by PERMANOVA (p = 0.016 and 0.041, respectively). Sex was significantly associated with both binary Jaccard and Bray-Curtis distances at the family level and below (p = 0.002 and 0.040, respectively). No significant associations were observed between alpha or beta diversity and prescribed psychotropic medications, STAI trait scores, or the number of DSM5 AUD criteria at screening. Smoking was only associated with a decrease in alpha diversity at the ASV level (ASV richness p = 0.044, Shannon diversity p = 0.027).

With respect to drinking measures, we observed significant alpha and beta diversity associations for both baseline drinks per week and PEth levels within our AUD cohort (Fig. 1b). Participants with higher screening PEth exhibited lower richness and Shannon diversity at the order level and below (p = 0.003 and < 0.001, respectively). Participants with higher baseline drinks per week exhibited lower richness at the order level and below (p < 0.001), but no significant change in Shannon diversity. Screening PEth and baseline drinks per week were significantly associated with binary Jaccard distances at the order level and below (p = 0.001 and 0.003, respectively). Additionally, screening PEth was significantly associated with Bray-Curtis distances below the genus level, while baseline drinks per week showed a significant association below the order level (p = 0.008 and 0.013, respectively).

After controlling for all individually significant variables in a PERMANOVA model, drinking measures remained significant and were the strongest predictors of genus-level community variance (Fig. 1c). This trend held across all distance metrics except Bray-Curtis distance and baseline drinks per week. Overall, higher alcohol exposure is associated with reduced microbiome alpha diversity and altered beta diversity, suggesting that drinking intensity remains an important determinant of gut microbiome structure among individuals diagnosed with AUD.

Given the role of age and sex in overall microbiome composition, we further examined the associations between baseline drinking behavior and microbiome alpha and beta diversity within age- and sex-stratified subgroups. To assess age effects, participants were divided into those older than the median age of 58 years (n = 60) and those 58 years or younger (n = 62). Participants were also stratified by sex into male (n = 76) and female (n = 46) groups. The demographic and clinical characteristics of the age- and sex- stratified subgroups are given in Supporting Tables 1 and 2. After adjusting for covariates, screening PEth was inversely associated with Shannon diversity across all subgroups at the genus or ASV levels (p = 0.002 in all samples, Fig. 2a). Screening PEth was only associated with a significant decrease in family and genus richness in the full cohort and younger subgroup (p = 0.029 and 0.028, respectively). Baseline drinks per week corresponded with a decrease in richness in the full cohort and in the younger and female subgroups from the family to ASV levels (p = 0.017, 0.021, and 0.013, respectively).

**Figure 2:**
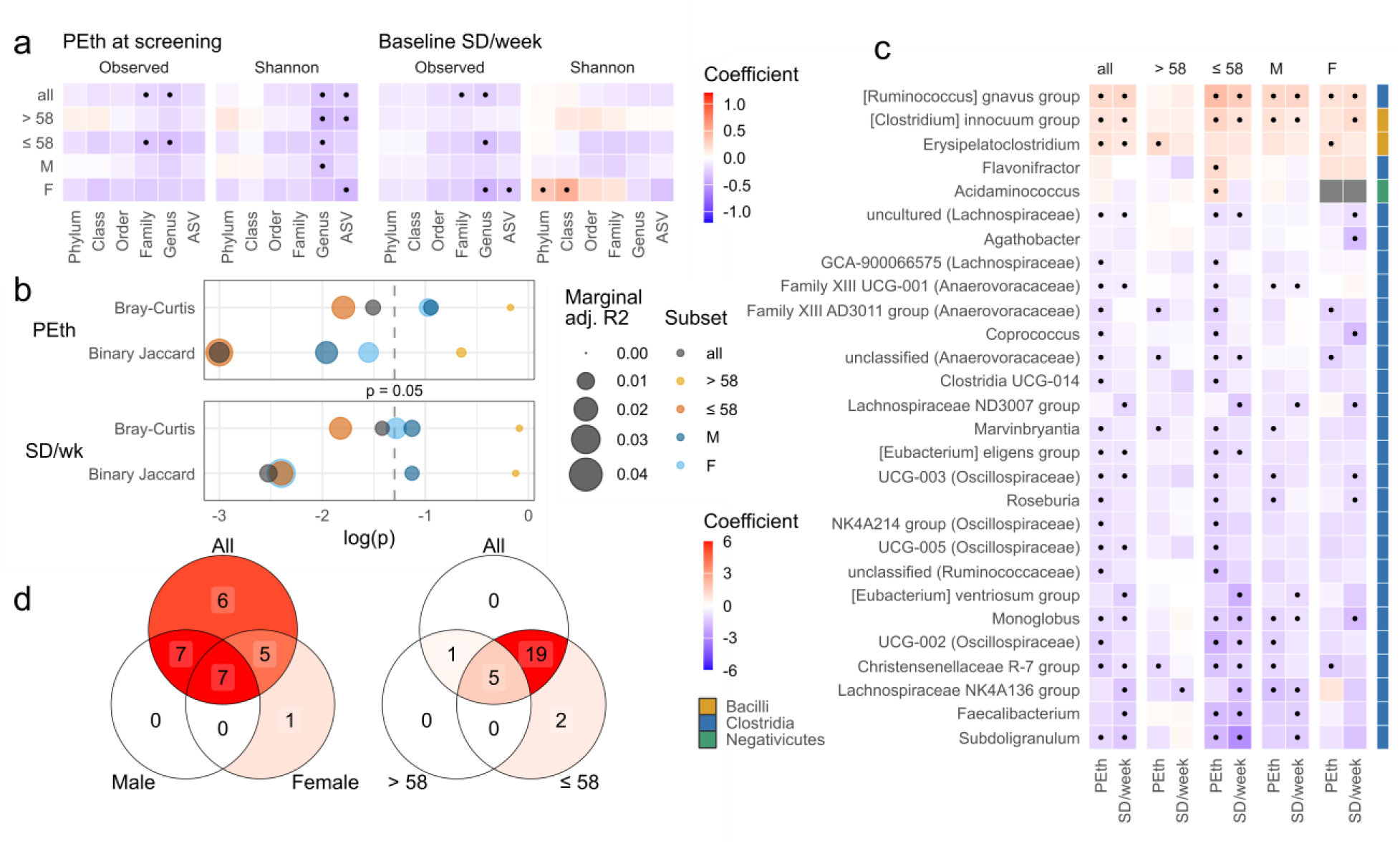
Microbiome changes associated with drinking. (a) Heatmap of alpha diversity associations with baseline PEth (left) and baseline drinks per week (right) controlled for covariates, split by subgroups. Dots in (a) and (c) indicate statistically significant associations (p < 0.05 and q < 0.1, respectively). (b) PERMANOVA R² values (controlled for covariates) for baseline PEth (top) and baseline drinks per week (bottom), split by subgroup. (c) Heatmap of associations between individual genera and drinking variables (x-axis), split by subgroup (plot facets). Color keys on the right indicate the taxonomic class of that genus. (d) Venn diagram showing the number of significant associations from Fig. 2c shared between all samples and the sex subgroups (left), and all samples and the age subgroups (right).

**Table 2:** AUC values for all participant and subgroup-specific models.

| Subgroup | Responder | Non-responder | Clinical Only | Metabolites Only | Microbiome Only | Clinical + Metabolites | Clinical + Microbiome | Clinical + Metabolites + Microbiome |
| --- | --- | --- | --- | --- | --- | --- | --- | --- |
| All participants | 77 | 21 | 0.60<br>(0.58, 0.62) | 0.78<br>(0.76, 0.81) | 0.71<br>(0.68, 0.74) | 0.77<br>(0.75, 0.80) | 0.73<br>(0.70, 0.75) | 0.76<br>(0.74, 0.79) |
| >58 years | 35 | 12 | 0.79<br>(0.75, 0.83) | 0.91<br>(0.89, 0.93) | 0.82<br>(0.79, 0.86) | 0.92<br>(0.90, 0.94) | 0.91<br>(0.88, 0.93) | 0.91<br>(0.88, 0.94) |
| ≤ 58 years | 42 | 9 | 0.82<br>(0.78, 0.85) | 0.90<br>(0.87, 0.93) | 0.84<br>(0.81, 0.87) | 0.88<br>(0.85, 0.91) | 0.85<br>(0.83, 0.88) | 0.94<br>(0.92, 0.96) |
| Male | 45 | 12 | 0.71<br>(0.68, 0.73) | 0.89<br>(0.87, 0.92) | 0.83<br>(0.80, 0.85) | 0.89<br>(0.87, 0.91) | 0.84<br>(0.81, 0.86) | 0.88<br>(0.86, 0.90) |
| Female | 32 | 9 | 0.73<br>(0.70, 0.76) | 0.91<br>(0.88, 0.94) | 0.89<br>(0.86, 0.92) | 0.89<br>(0.86, 0.92) | 0.90<br>(0.87, 0.93) | 0.92<br>(0.89, 0.94) |

In terms of beta diversity, the younger subgroup showed significant associations with drinking measures across distance metrics, while the older subgroup did not (Fig. 2b). Sex-stratified associations varied by metric: although neither sex subgroup exhibited a relationship with Bray-Curtis distances, binary Jaccard distances were significantly associated with PEth in both groups and with drinks per week in the female subgroup. Furthermore, the female subgroup generally had higher PERMANOVA R² values than the male subgroup, except in the case of screening PEth and binary Jaccard distances. No significant sex differences were observed across the age subgroups, and no significant age differences were observed across the sex groups (Supporting Table 1, 2). These findings suggest that the relationship between drinking measures and gut microbiome community structure is modified by age and sex, with consistently stronger associations in younger participants.

### Specific Taxa at Baseline Are Associated with Drinking Intensity

The community-level relationship between baseline drinking characteristics and microbiome composition was further confirmed by analysis of individual taxa. Using GLM, we identified 25 genera associated with either drinking measure in the full cohort. Individual genera correlated with age, sex, and BMI in the same GLM model are shown in Supporting Fig. 2. Four genera were only associated with baseline drinks per week, 10 were only associated with screening PEth, and 11 were associated with both (Fig. 2c). Three additional genera were associated with drinking in only the younger or female subgroups. Most identified genera were members of the class *Clostridia*, suggesting that *Clostridia* serves as a hub taxon for microbiome changes related to drinking intensity. Additionally, most microbe–drinking associations were negative. Only three genera, [*Ruminococcus*] *gnavus* group, [*Clostridium*] *inocuum* group and *Erysipelatoclostridium*, were positively correlated with baseline drinking across both measures.

Subgroup analyses support the earlier finding that alcohol-associated microbial populations are influenced by age and sex. Participants in the >58-year-old group showed fewer significant (p < 0.05) associations, with only five genera negatively correlated and one genus positively correlated with either drinking measure (Fig. 2c, 2d). In contrast, participants ≤58 years old exhibited a larger number of significant associations that overlapped more strongly with those identified in the full cohort (Fig. 2c, 2d). Two genera, *Flavonifractor* and *Acidaminococcus*, were uniquely correlated with screening PEth in the younger cohort (Fig. 2c, 2d). Additionally, *Erysipelatoclostridium* was significant in the older but not the younger group. Overall, these results suggest that the observed associations between the gut microbiome and drinking intensity in the full cohort are primarily driven by shifts in younger participants.

The divergence in microbiome change was less pronounced between the sex subgroups. Out of the 25 genera significantly related to screening PEth levels and/or baseline drinks per week, 7 were shared by both male and female participants, 5 by only female participants, and 7 by only male participants (Fig. 2d). *Agathobacter* was the only genus unique to the female subgroup that was not also associated with drinking in the male subgroup or entire cohort (Fig. 2c).

### Baseline Gut Metabolites are Associated with Drinking Intensity

After quality filtering, 159 out of 356 metabolites from the targeted panel were evaluated in the metabolome analysis. The overall stool metabolome was significantly associated with age, sex, and both drinking measures when testing individual variables by PERMANOVA (Fig. 3a), consistent with the microbiome beta diversity results (Fig. 1b). Similarly, after accounting for age and sex in the PERMANOVA model, screening PEth and drinks per week remained the strongest predictors of metabolome variance (Fig. 3b). PERMANOVA analysis within age and sex subgroups showed that the effect of drinking on the overall metabolome was maintained across most strata, except in the older subgroup for baseline drinks per week and in the female subgroup for screening PEth (Fig. 3c).

**Figure 3:**
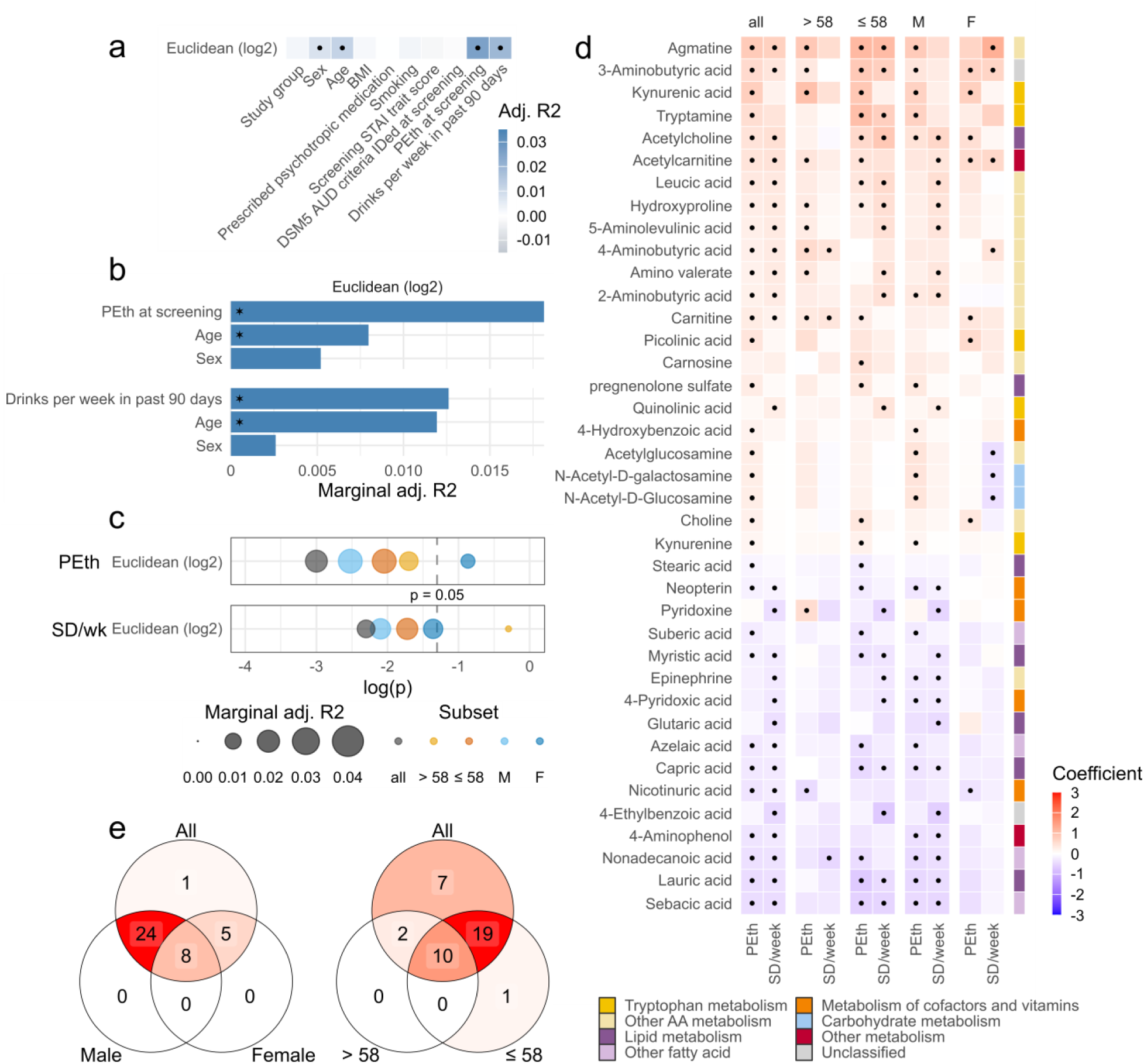
Metabolome changes associated with drinking. (a) Heatmap of single-variable PERMANOVA tests for metabolome samples. Dots in (a) and (d) indicate statistically significant associations (p < 0.05 and q < 0.1, respectively). (b) Bar plot of PERMANOVA R² values (controlled for covariates) at genus level for clinical variables significantly associated with the single-variable analysis. (c) PERMANOVA R² values (controlled for covariates) for baseline PEth (top) and baseline drinks per week (bottom), split by subgroup. (d) Heatmap of associations between individual metabolite features and drinking variables (x-axis), split by subgroup (plot facets). Color keys on the right indicate the KEGG pathway classification of that metabolite. (e) Venn diagram showing the number of significant associations from Fig. 3c shared between all samples and the sex subgroups (left), and all samples and the age subgroups (right).

Analysis of specific metabolites using GLM identified 38 gut metabolites correlated with drinking intensity in the full cohort: 6 were only associated with baseline drinks per week, 12 were only associated with screening PEth, and 20 were associated with both (Fig. 3d). Individual metabolites correlated with age, sex, and BMI in the same GLM model are shown in Supporting Fig. 2. In general, gut metabolites involved in amino acid metabolism were positively associated with drinking, whereas metabolites involved in lipid metabolism (largely medium-chain saturated fatty acids) and the metabolism of cofactors and vitamins were negatively associated with drinking. Notably, drinking intensity was positively associated with a number of metabolites (tryptamine, kynurenic acid, picolinic acid, and kynurenine) involved in the kynurenine pathway of tryptophan metabolism.

Similar to the pattern observed in the drinking-associated microbiota, the >58-year-old group showed fewer significant associations between baseline drinking and metabolites compared to the ≤58-year-old group (Fig. 3d, e). Unlike the microbiome results, however, this weakened relationship was also observed within the female subgroup. The negative associations between drinking intensity and lipid and cofactor metabolism observed in the younger and male subgroups were distinctly lacking in the older and female subgroups. However, the positive associations between drinking intensity and metabolites involved in amino acid metabolism remained mostly evident across all subgroups.

In summary, the metabolome findings suggest that metabolites associated with amino acid metabolism increased with drinking intensity, whereas those associated with lipid metabolism decreased. Age and sex substantially modify the relationship between drinking intensity and the stool metabolome, with a more evident decrease in stool lipids among male participants and younger participants. Female participants and older participants exhibit a more selective metabolic response, particularly involving amino acid metabolism.

### Co-variation of the Gut Microbiome and Metabolome with Drinking Intensity

Given that the gut microbiome plays an important role in shaping the gut metabolome, we sought to identify metabolites that were strongly associated with bacterial abundances in our AUD cohort. Specifically, we used GLM to correlate the abundance of microbial taxa and metabolites that were significantly associated with either drinking measure (Fig. 4, Supporting Fig. 3). Microbial taxa positively associated with drinking (Fig. 4a, numbers 1-5) covaried with metabolites involved in amino acid metabolism and were inversely related to lipid metabolites. Microbes negatively associated with drinking (Fig. 4b, numbers 6-24) exhibited the opposite pattern, co-varying with lipid metabolites and inversely varying with amino acid metabolites. The consistency of this relationship across metabolites and taxa suggests that compositional changes related to higher drinking intensity are accompanied by a coordinated functional shift favoring amino acid metabolism over lipid metabolism.

**Figure 4:**
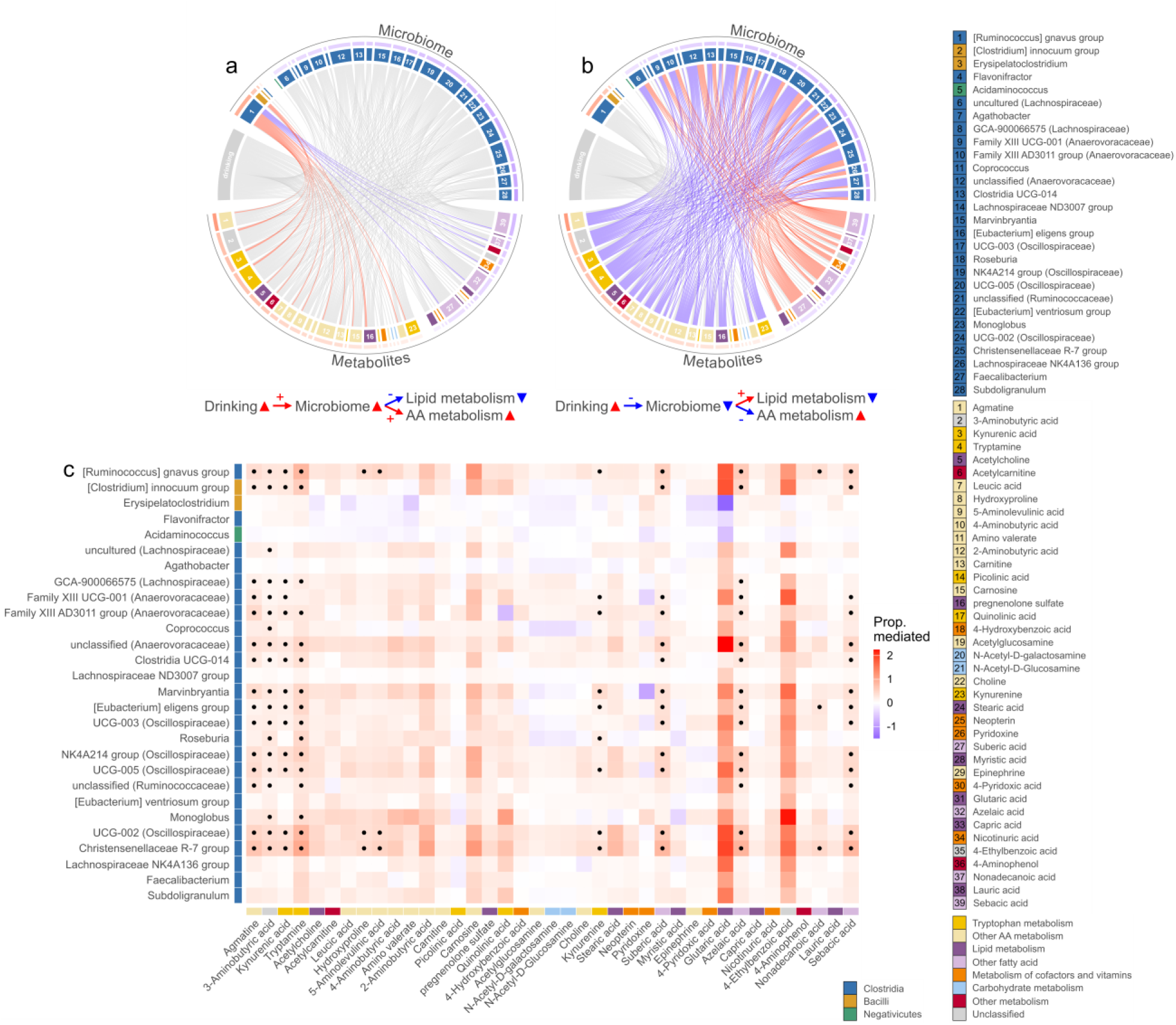
Microbiome-metabolome co-associations. (a/b) Chord diagram showing significant associations between microbiome and metabolites for microbiome genera that significantly increase in abundance (a) and decrease in abundance (b) with drinking. Each arc inside the ring shows one significant association between a microbiome and metabolite feature (q < 0.01). Red and blue arcs indicate a positive and negative co-association, respectively. Grey arcs in each plot are either co-associations that will be highlighted in the other plot or represent the association that every feature has with drinking. If a microbiome or metabolite feature has no co-associations with the other type of feature, it is shown with only a single arc connecting to the “drinking” variable on the outer ring. The outer ring shows what two features are involved in a co-association. Microbiome genera (top, 1–28) and metabolites (bottom, 1–39) are labeled numerically in the same order as Figures 2 and 3. The small bar above each microbiome and metabolite feature on the outer ring reflects the average coefficient of that feature across baseline PEth and drinks per week in the full cohort, with the same red/blue color scale described above. (c) Heatmap showing mediation analysis results testing for the mediating effect of microbiome features on the PEth-metabolite relationship. The x-axis represents the metabolite included in the mediation analysis model, and the y-axis represents the microbiome feature included in the mediation analysis model. Dots indicate a statistically significant mediation effect (q < 0.1). Color keys in the chord diagram and outside the heatmap indicate the KEGG pathway classification of each metabolite and taxonomic class of each microbiome genus.

To further elucidate the relationship between metabolites, the microbiome, and drinking intensity, we performed mediation analysis testing for the mediating effect of microbiome features on the correlation between screening PEth and metabolites, adjusted for covariates (Fig. 4c). Most drinking-associated genera were mediators for dicarboxylic fatty acids (suberic acid, azelaic acid, and sebacic acid), which are products of omega-oxidation of fatty acids, and tryptophan metabolites (kynurenic acid, tryptamine, and kynurenine). The gut microbiome has been reported to modulate host tryptophan metabolism and tryptophan metabolite levels in the colon.^30, 34^ Agmatine, an intermediate in polyamine biosynthesis, and 3-aminobutyric acid (BABA) were also strongly mediated by microbiome features. For these metabolites, the mediating effect of microbiome features was consistently observed across both positively and negatively drinking-associated taxa. Identical mediation effects were observed when metabolites were modeled as mediators. Although a definitive causal flow for these relationships cannot be established without longitudinal data, this result suggests that some drinking-associated metabolites and microbial taxa exhibit a tightly coupled and potentially bidirectional relationship.

### Baseline Omics Improves Classification Models for Drinking Reduction

Although clinical data have yielded modest success in classifying AUD treatment outcomes, baseline microbiome and metabolome profiles may capture additional biological factors that explain predisposition to treatment response.^35, 36^ To explore this, we applied a random forest classification approach on each dataset alone and in combination with the other datasets. Endpoint drinking data were available for 98 out of the 122 subjects with baseline stool samples. Participants who exhibited a clinically meaningful drinking reduction were classified as responders (n = 21), and those who did not were classified as non-responders (n = 77, demographic information in Supporting Table 3). Random forest models were constructed using single or combined sets of clinical, metabolite, and/or microbiome data at baseline to classify responders and non-responders after dutasteride or placebo treatment. Notably, metabolite data appeared to have the largest impact on model performance. The metabolite-only classifier (AUC = 0.78, 95% CI: 0.76–0.81) outperformed the clinical or microbiome data-only dataset (clinical AUC = 0.60, 95% CI: 0.58–0.62, microbiome AUC = 0.71, 95% CI: 0.68–0.74) and exhibited comparable performance to the classifier integrating all three datasets (AUC = 0.76, 95% CI: 0.74–0.79, Fig. 5a, Table 2).

**Figure 5:**
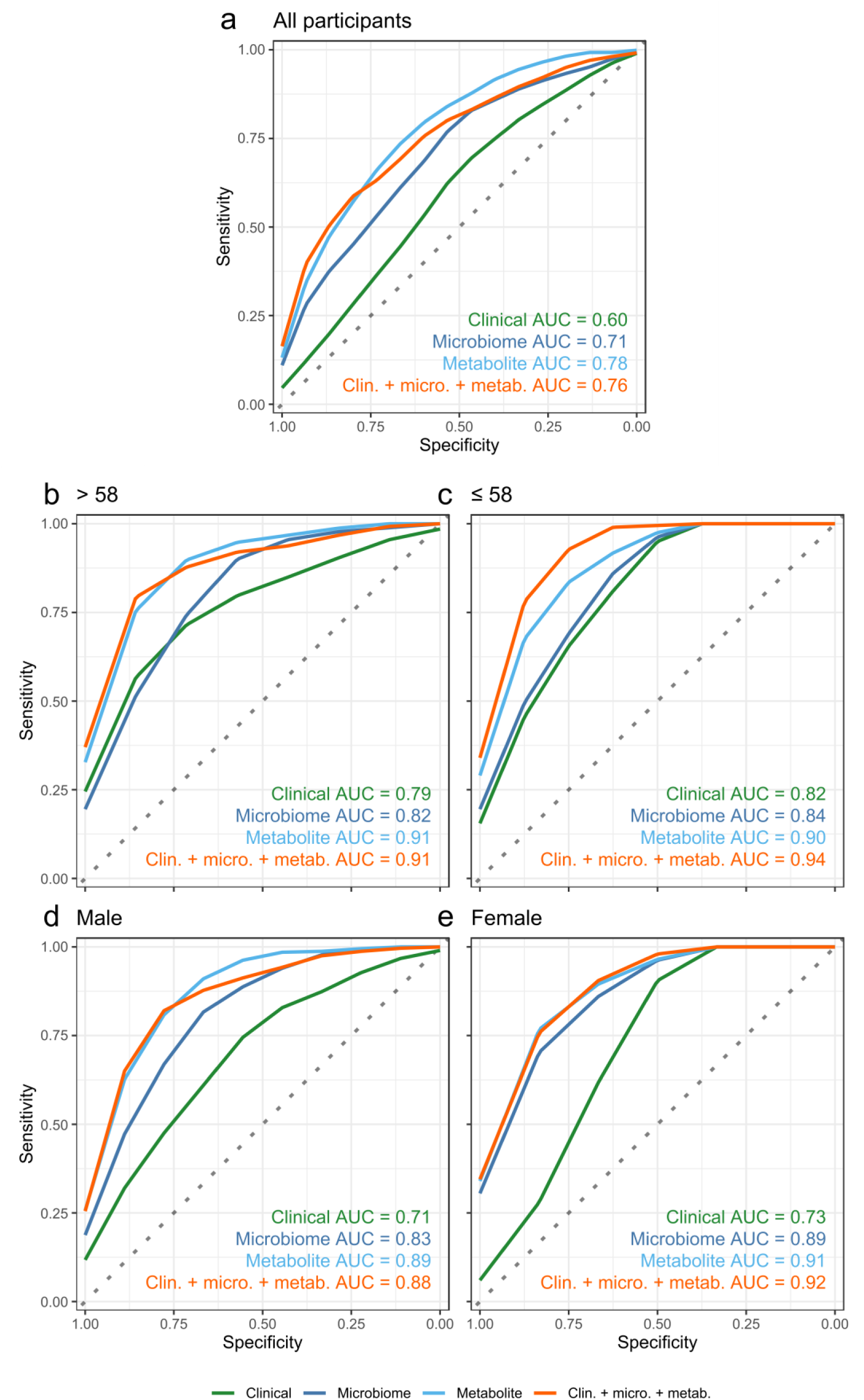
Random forest classifier performance for drinking responders vs. non-responders. (a-e) Receiver Operating Characteristic (ROC) curves for random forest classification models constructed using all participants (a), participants over 58 years old (b), participants 58 years and younger (c), male participants (d), and female participants (e). For each subset, AUC values for each model type are shown at the bottom right of the plot area.

Additionally, microbiome data alone or in combination with other information led to a smaller but notable classification improvement over clinical data alone (Supporting Fig. 4). Supporting Fig. 6 shows feature importance for all models.

**Figure 6:**
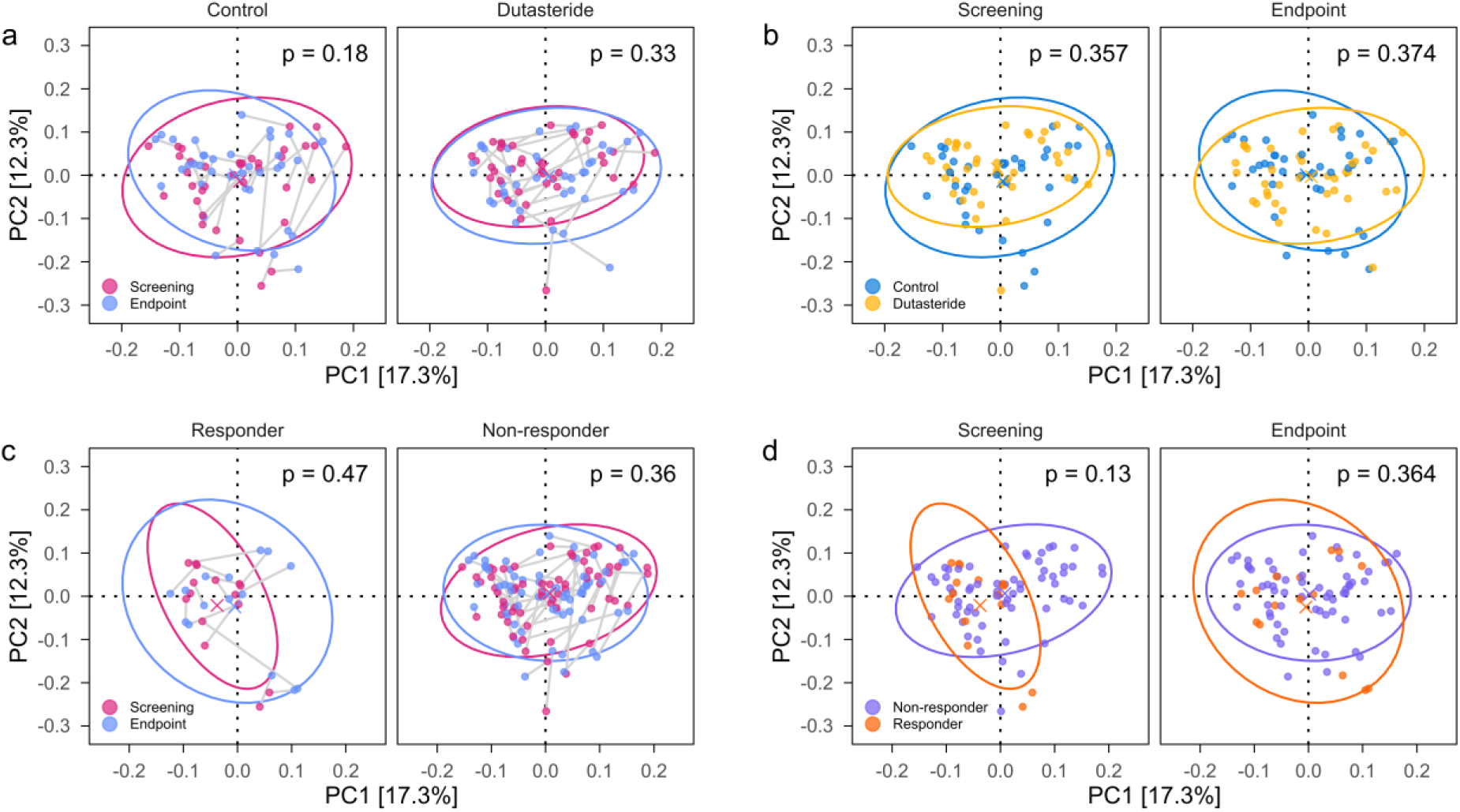
Microbiome changes due to drug treatment and intervention response. (a/c) PCoA plot and PERMANOVA results of paired stool samples testing for the difference between timepoints between (a) control and dutasteride participants and (c) drinking responder and non-responder participants (defined as a <60% reduction from baseline drinks per week). (b/d) PCoA plot and PERMANOVA results of paired stool samples testing for the difference between (b) treatment groups and (d) drinking responders and non-responders for participants at screening and endpoint. All analyses were performed using Bray-Curtis distances at the genus level.

Given the age- and sex-dependent relationships between drinking and both microbiome and metabolites, we also stratified models by age and sex. Although the smaller sample size of responders limits definitive interpretation, the stratified classifiers followed a pattern similar to the full cohort, where model performance increased when trained on -omics data (Fig. 5b, 5c, Table 2). Age- and sex-stratified feature importance plots are shown in Supporting Figs. 7 and 8.

Overall, classifier performance improved with the integration of multiple datasets, and distinct demographic effects were observed with age. Metabolite data exhibited the most consistent increase in classification ability compared to microbiome data or clinical data.

### Dutasteride Treatment and Drinking Reduction Have Minimal Impact on the Gut Microbiome

To investigate the impact of dutasteride treatment on the gut microbiome, we compared the microbial community in participants with sequenced samples at both time points (n = 77). This subset exhibited comparable demographics to the baseline-only cohort, with the exception of a significantly lower BMI (p = 0.041, Supporting Table 4). PERMANOVA revealed no significant difference in microbiome composition from baseline to endpoint within either treatment group (Fig. 6a; demographic information in Supporting Table 5). Additionally, between-group comparisons at either baseline or endpoint did not show a significant difference between treatment groups (Fig. 6b).

Given that not all participants met the threshold for a clinically meaningful response (defined as a >60% decrease in drinks per week) after dutasteride treatment, we also tested whether drinking reduction was associated with a change in microbiome over the course of the study. Participants who exhibited a clinically meaningful drinking reduction were classified as responders (n = 15), and those who did not were classified as non-responders (n = 62, demographic information in Supporting Table 6). No significant microbiome shifts were observed for within-group (Fig. 6c) and between-group (Fig. 6d) comparisons using PERMANOVA. Neither dutasteride treatment nor drinking response were associated with significant changes in alpha diversity. Linear mixed regression analysis also did not identify any individual taxa that changed after dutasteride treatment or drinking response after FDR correction.

## Discussion

Leveraging a large, randomized intervention trial in patients with AUD,^1^ we show that drinking intensity remains an important determinant of gut microbiome diversity and composition even within a clinically defined AUD population. The dose-dependent nature of AUD-associated microbiome changes likely explains inconsistent findings of taxa associated with drinking in previous case-control studies.^11, 29, 44^ Our data indicates that heavier drinking is associated with a general decline in microbiome diversity and decreased abundance of a broad range of taxa in the class *Clostridia* (specifically various *Lachnospiraceae*, *Oscillospiraceae*, and *Anaerovoraceae*), many of which have been implicated in butyrate fermentation.^45–47^ This result aligns with prior reports of depleted SCFA levels in individuals with higher alcohol consumption.^9, 48, 49^

While most drinking-microbiome associations were negative, a few microbes increased in abundance in the AUD population, most notably *Ruminococcus gnavus*. Although *R. gnavus* is a common constituent of the human gut flora, it is consistently enriched in inflammatory conditions such as AUD, fatty liver disease, and inflammatory bowel disease (IBD).^50, 51^ The observed expansion of *R. gnavus* is likely driven by several competitive advantages in the inflamed gut, including reduced competition, adaptation to oxidative stress, and, as a mucin glycan forager, the ability to exploit higher mucus turnover during inflammation.^52, 53^ Some strains of *R. gnavus* may further exacerbate inflammation by triggering cytokine production in response to their capsule polysaccharides.^54^ Other microbes enriched in our cohort include *Erysipelatoclostridium* and *Clostridium innocuum,* which have been implicated in IBD.^55, 56^ These microbes may represent potential therapeutic targets for addressing AUD-associated inflammation and gut barrier dysfunction.

Previous work on the metabolome in alcohol consumption has largely focused on circulating metabolites.^57^ Our analysis of stool metabolites shows broadly positive associations between drinking intensity and amino acid intermediates, particularly those involved in the kynurenine pathway of tryptophan metabolism. Gut tryptophan metabolism is strongly shaped by microbial activity, and has been implicated in craving, relapse, and psychological state in AUD.^30, 58^ Fecal kynurenine, neuroprotective kynurenic acid, and neurotoxic picolinic and quinolinic acid are all positively associated with drinking intensity in our cohort, which may implicate microbial metabolism in AUD-associated neurochemical dysfunction. The observed increase of both neurotoxic and neuroprotective metabolites, rather than changes in opposite directions, likely reflects an increase in amino acid metabolism in the gut. This contrasts with studies of circulating kynurenine pathway metabolites, which typically describe an increase in quinolinic acid and a decrease in kynurenic acid.^30, 59^ Further examination of this discrepancy between stool and circulating levels and their relationship with neuroactivity may offer novel insights into the contributions of gut metabolism to AUD pathology.

Tryptamine and agmatine were also positively associated with drinking intensity and mediated by microbiota. Tryptamine stimulates gut motility and serotonin release, while agmatine is a neuroprotective arginine metabolite that also shows anti-inflammatory effects through the inhibition of nitric oxide synthase (NOS).^60^ *R. gnavus* may therefore be particularly relevant to gut-brain communication due to its capacity to metabolize tryptophan to tryptamine, as well as arginine to agmatine.^61^ *R. gnavus* and *Erysipelatoclostridium ramosum* have been shown to be implicated in gut serotonin release in vivo.^62^ It has also been shown that NOS synthase activity inhibits *R. gnavus* growth in vivo, raising the possibility that the observed stool agmatine levels reflect a response by *R. gnavus* populations to host immunity.^61^ Overall, our microbiome-metabolome data suggest that microbial taxa enriched in AUD exert a mix of stimulatory and inhibitory effects on enteric immunity and nervous signaling. The role of these features, particularly *R. gnavus*, in inflammation and the gut-brain axis of AUD patients warrants continued investigation.

We observed a negative association between stool lipid levels and drinking intensity, which aligns with similar findings in human and primate studies of AUD and fatty liver disease.^63^ Mediation analysis suggests a link between microbiota and medium-chain dicarboxylic acids (MCDAs), which are produced by omega oxidation of fatty acids. Alcohol has complex effects on hepatic lipid metabolism and is known to inhibit beta oxidation in the liver, thereby shifting fatty acid metabolism toward omega oxidation. It has been shown that upregulation of omega oxidation can manifest as higher levels of MCDAs in stool.^64^ The observed decline may suggest that this effect is overwhelmed by increased systemic retention that limits substrates for microbial lipid metabolism, or by a depletion of lipid-metabolizing microbes due to AUD-associated disruption. Given that microbiome changes due to alcohol consumption have been restored by feeding saturated long-chain fatty acids to mice, modulation of lipid metabolism may be an interesting target for future work.^65^

A unique contribution of this study is the finding that age and sex modify the relationship between drinking and the gut microbiome and metabolome. In the male and younger subgroups, we observe robust associations with amino acid and lipid metabolism, which are limited to a few amino acid metabolites in the female and older subgroups. Aging is characterized by increased individualization and a pro-inflammatory shift in the microbiome and metabolome.^66, 67^ In our cohort, older participants and female participants also reported lower average alcohol consumption than younger participants and male participants, respectively (Supporting Table 1, 2). Thus, the absence of significant microbiome-drinking correlations in older participants may be explained by higher baseline inflammation, increased heterogeneity that obscures uniform trends, or reduced alcohol consumption. The observed sex differences may similarly be attributed to lower alcohol exposure or to sex-dependent differences in lipid metabolism that buffer against alcohol-induced metabolic perturbations.^68^ These findings are particularly relevant as AUD phenotypes differ by age and sex,^17–25^ underscoring the need for demographically-informed evaluation and treatment strategies.

In addition, we examined whether a reduction in drinking would influence microbiome composition. A prior small-scale study (n = 22) reported microbiome changes over 4 weeks following abstinence in AUD, and another cohort study (n = 63) observed an increased abundance of butyrate fermenters and a reduction in *R. gnavus* among patients who maintained abstinence after 14 days of withdrawal treatment.^12, 70^ These mirror the patterns we observed with lower alcohol intake in our baseline data. However, we did not observe a significant shift in microbial community structure when comparing baseline to endpoint. This may be attributable to the modest average reduction in drinking (31%) observed over the 12-week study period. It may be possible that recovery from AUD-associated microbiome disruption requires a sustained reduction in drinking, which we did not observe among most patients in our study (Supporting Table 5).

One important direction in advancing precision treatment approaches for AUD is to identify individuals who are more likely to respond to pharmacological and behavioral treatment. Using our comprehensive clinical, microbiome, and metabolome data, we constructed random forest classifier models for endpoint treatment response in the full cohort as well as subgroups stratified by sex and by age. Classifier performance consistently improved when clinical data was augmented with both microbiome and metabolite data, suggesting that microbiome and metabolome profiles contain relevant signals for endpoint treatment response in our cohort. To rule out cohort-specific effects and validate the clinical utility of multi-omic signatures as a predictive tool for AUD treatment, it will be necessary to investigate whether this finding is generalizable across broader studies of AUD patients.

This study has several limitations. The inclusion of detailed dietary records and antibiotic and complete medication histories would provide more clarity on additional confounding variables. Due to the limited availability of sample material, we were unable to obtain stool metabolomic profiles at the study endpoint. Follow-up research using animal models will be necessary to validate the proposed mechanistic links between the identified biological markers and drinking behavior. Additionally, longitudinal -omics data from human cohorts could help establish directional causation beyond the associations found in our cross-sectional mediation analysis.

Overall, our findings reveal complex, demographic-dependent associations between the gut microbiome and metabolome, drinking intensity, and treatment response in AUD. These results enhance our understanding of the microbiome in AUD and provide a foundation for future precision medicine strategies aimed at tailoring interventions to individualized microbial and metabolic signatures.

## Methods

### Human Trial

This study was conducted with approval from the UConn Health IRB in accordance with relevant guidelines and regulations. Participants were recruited from the community as part of a double-blind, randomized, placebo-controlled study investigating the use of dutasteride in reducing drinking (ClinicalTrials.gov identifier: NCT04098302, Dutasteride Treatment for Reducing Heavy Drinking in AUD: Predictors of Efficacy, first registered 19/09/2019). The study protocol is published online.^1^ Eligible participants were aged 35-70 and regularly consumed >24 (male) or >18 (female) standard drinks per week, with at least 2 days per week classified as heavy drinking days (≥5 drinks per day for male participants, ≥4 drinks per day for female participants). Participants were also required to meet DSM-5 diagnostic criteria for alcohol use disorder and express a desire to stop drinking or reduce their drinking to non-hazardous levels. Female participants of child-bearing age were required to practice a reliable method of birth control and have a negative serum pregnancy test result prior to study initiation. Potential participants who were currently lactating, exhibited significant cognitive impairment, physical disease, or psychiatric illness, had a history of serious alcohol withdrawal symptoms, concurrent DSM-5 diagnosis of a drug use disorder (other than caffeine or nicotine), or taking finasteride, dutasteride, other medication for treatment of AUD, or opioid pain medication were excluded. Participants were assigned treatment groups via urn randomization, balancing for AUD severity based on DSM-5 criteria, current smoking status, and use of psychiatric medication. Separate randomizations were conducted for men and women to ensure balance in both sex groups.

The treatment group received a daily dose of 1 mg dutasteride for 12 weeks. Bi-weekly medical management was provided for both groups. Drinking at baseline (over the past 90 days) and endpoint (over the last 4 weeks of the study) was estimated using the timeline follow-back method (TLFB). The trait version of the Spielberger State-Trait Anxiety Inventory (STAI) was administered at baseline.

### 16S rRNA Sequencing and Data Processing of Stool Samples

Stools were collected by participants and stored on ice up to 24 hours prior to baseline and 12-week (endpoint) visits. Stool aliquots were prepared upon receipt at the clinical research center at UConn Health and stored at -80 ℃ until the time of analysis. Microbial DNA was isolated from stool samples using the PowerSoil DNA Isolation kit (Qiagen) following the manufacturer’s instructions. Bacterial 16S ribosomal RNA (rRNA) gene sequencing was performed on V4 hypervariable regions using using 515F (5’- GTGYCAGCMGCCGCGGTAA-3’) and 806R (5’- GGACTACNVGGGTWTCTAAT-3’) primers. The resulting amplicon libraries were then purified using Zymo Select-a-Size MagBeads (Zymo), quantified (Qubit 2.0 fluorimeter, Invitrogen), and pooled with equal masses added from each sample. The initial pool underwent two additional cleanup steps using Zymo Select-a-Size MagBeads (Zymo). The pooled and purified library was sequenced on the Illumina MiSeq platform (Illumina) using 2 × 250 bp, 500 cycles kits.

Raw 16S rRNA sequencing data was basecalled with Illumina RTA (v1.18.54.4) and converted into demultiplexed FASTQ files using bcl2fastq2 (v2.20). The FASTQ output was subsequently processed using QIIME2 (v2023.09).^71^ First, reads were denoised using the dada2 plugin in QIIME2 with options “denoise-paired --p-trim-left-r 0 --p-trunc-len-f 250 --p-trunc-len-r 250 --p-trim-left-f 0 --p-n-threads 0.” Taxonomic classifications were assigned to the resulting amplicon sequence variants (ASVs) with the QIIME2 feature-classifier plugin using a pre-trained Naive Bayes classifier based on the SILVA 138 99% OTU database.^72–74^ The feature-classifier plugin was executed with the following options: “classify-sklearn --i-classifier silva-138-99-nb-classifier.qza --p-n-jobs -3.” The output matrix of ASV counts was then normalized by sequencing run to minimize inter-run variation using ComBat() from the sva package in R (v3.56.0).^75^

### Targeted LC-MS Analysis of Stool Metabolome

A panel of 356 metabolites representing a diverse set of metabolic pathways were analyzed using a targeted LC-MS approach, as described previously.^76–78^ Approximately 20 mg of stool from each sample was suspended in 200 μL 4:1 methanol:phosphate buffered saline containing 1.8104 mM ^13^C_3_-lactate and 142 μM ^13^C_5_-glutamic acid and homogenized by bead beating using 400 μM zirconium grinding media (OPS Diagnostics). Homogenate was quenched with 800 μL 4:1 methanol:phosphate buffered saline containing 1.8104 mM ^13^C_3_-lactate and 142 μM ^13^C_5_-glutamic acid, vortexed thoroughly, and incubated 30 min at -20℃. Samples were then sonicated for 30 min in an ice bath and pelleted for 10 min at 14,000 RPM and 4℃. 800 μL of supernatant was transferred to a new microcentrifuge tube and dried under vacuum in a CentriVap Concentrator (Labconco). Dried residue was resuspended in 150 μL 40% phosphate-buffered saline/60% acetonitrile. A quality control sample was prepared by pooling an equal portion from all prepared samples.

All LC-MS analysis was performed on an Agilent 1290-UPLC-6490 QQQ-MS instrument with an electrospray ionization source controlled using Agilent MassHunter Workstation software. Each sample was injected twice, 10 μL using negative ionization mode and 4 μL using positive ionization mode. Both separations were performed using hydrophilic interaction chromatography on a Waters XBridge BEH Amide column (150 x 2.1 mm, 2.5 μm particle size). The flow rate was kept at 0.3 mL/min, auto-sampler at 4℃, and column compartment at 40℃. The mobile phase consisted of solvent A (10 mM ammonium acetate, 10 mM ammonium hydroxide in 95% deionized water/5% acetonitrile) and solvent B (10 mM ammonium acetate, 10 mM ammonium hydroxide, 5% deionized water/95% acetonitrile). After 1 min isocratic elution at 90% B, the mobile phase ramped to 40% B over 10 min, maintained at 40% B for 4 min, and finally ramped to 90% B to prepare for the next injection. Targeted data acquisition was performed in multiple-reaction-monitoring mode. The extracted multiple-reaction-monitoring peaks were integrated using Agilent MassHunter Quantitative Data Analysis software. Of the 356 analyzed metabolites, 159 metabolites met the QC criteria for inclusion (CV <20% across quality control pools and present in >80% of all samples) and were retained for downstream analysis in R. The peak heights for these metabolites were normalized for sample mass by dividing each peak height in a sample by the grams of stool used for metabolomics.

### Statistical Analysis

Statistical analysis was performed in R (v4.5.0). Clinical variables within the stool sample cohort were assessed for significant differences using the gtsummary package (v2.2.0).^79^ Fisher’s exact test was applied to categorical variables and Welch’s two-sample t-test was applied to continuous variables.

For alpha and beta diversity analyses, samples were first normalized for read depth by down-sampling to 10,000 reads per sample before calculating the measurement. Alpha diversity metrics were calculated using estimate_richness() from phyloseq (v1.52.0)^80^, and beta diversity metrics were calculated using vegdist() and pco() from vegan (v2.6.10)^81^. Alpha diversity was fitted to a linear model against Z-scored clinical data using stats::lm() and base::scale() in R (v.4.5.0). All beta diversity associations were assessed for significance using permutational multivariate analysis of variance (PERMANOVA) as implemented in adonis2() from vegan (v2.6.10). PERMANOVA effect sizes were adjusted to account for varying degrees of freedom (omega squared, ω²) as implemented in adonis OmegaSq() from the MicEco package (v0.10.0).^82^ Alpha diversity and PERMANOVA results in Figures 1c, 2a, 2b, 3b, and 3c included the drinking variable as well as sex, BMI, age, psychotropic medications, smoking, and STAI trait score as co-variates. For all alpha and beta diversity analyses, p < 0.05 was considered significant.

To assess relationships between individual microbiome or metabolite features and the clinical data, feature abundances were fitted to a generalized linear model using the Maaslin2 package (v1.22.0).^83^ In brief, the bacterial genera and metabolites data were filtered to retain features present in > 25% of samples, then each feature was log2-transformed and used as the response variable for a generalized linear model. Predictor variables for the model included one of the two drinking variables (screening Peth or baseline drinks per week), as well as the following baseline covariates: sex, BMI, age, psychotropic medication use, smoking status, and STAI trait score. To correct for multiple comparisons, p-values within each analysis were adjusted using the Benjamini–Hochberg false discovery rate correction as implemented in stats::p.adjust(method = “BH”) (v4.5.0). Features with a q < 0.10 were considered significant.

Significant metabolites after filtering were functionally classified using Human Metabolome Database (HMDB, v5.0) ^84^ annotations as follows: HMDB IDs were converted into Kyoto Encyclopedia of Genes and Genomes (KEGG)^85^ compound IDs, which were then matched to the KEGG pathways corresponding to each compound ID. The best pathway match was manually annotated with a broader functional class based on the KEGG BRITE hierarchy database.

Co-association analyses between microbial and metabolite features were similarly performed by fitting log2-transformed abundances to a generalized linear model using stats::glm() (v4.5.0). For microbe-metabolite associations, microbial genus abundance was modeled as the response variable against metabolite abundance as the predictor variable. A total of 16,695 such tests were conducted across 105 genera and 159 metabolites. To correct for multiple comparisons, p-values for microbe-metabolite associations were adjusted using the Benjamini & Hochberg method and a more conservative q < 0.01 threshold was applied.

To evaluate whether gut microbiome features mediate the relationship between screening PEth and metabolite features, bootstrapped mediation analysis was conducted using the mediation package in R (v4.5.1)^86^ for 500 iterations. The independent variable for the model was set to screening PEth, the mediator set to gut microbiome, and the outcome variable was set to metabolite for each combination of drinking-related microbiome and metabolite features. We then swapped the mediator and outcome variable to evaluate which metabolites mediated the relationship between screening PEth and a given microbiome feature. Models were controlled for sex, BMI, age, psychotropic medications, smoking, and STAI trait score. For each set of mediation analyses, a total of 1,092 tests were conducted across 28 genera and 39 metabolites. P-values were FDR-adjusted as implemented in stats::p.adjust(method = “BH”) (v4.5.0), and features with a q < 0.10 were considered significant.

Plots were generated in R 4.5.0 using packages ggplot2 (v3.5.2)^87^, ggpubr (v0.6.0)^88^, ggh4x (v0.3.0)^89^, circlize (v0.4.16)^90^, rstatix (v0.7.3),^91^ microViz (v0.12.3)^92^, and Polychrome (v1.5.4)^93^.

### Random Forest Predictive Model

Random forest was used to classify whether participants experienced a clinically relevant drinking reduction after treatment (>60% reduction in drinking). All available baseline clinical and demographic variables were included as input features in the clinical models (treatment group, sex, age, BMI, height, weight, psychotropic medication use, smoking status, primary race, screening STAI trait score, DSM5 AUD criteria identified at screening, average drinks per week at screening, average heavy drinking days per week at screening, and screening PEth level). Microbiome models included relative abundances of all features at baseline that were present in > 25% of samples, and metabolome models included peak heights normalized for sample mass as described previously. First, we performed feature selection using the random forest–based recursive feature elimination (RFE) method (caret, v7.0.1).^94^ To optimize feature selection, this procedure was repeated 100 times. The input data was filtered to only include the consensus feature set, defined as the features that appeared in more than 50 RFE iterations.

After determining the final feature set, model training was conducted. Five different datasets were used: all participants (responders: 21; non-responders: 77), and subgroup analysis by age and sex (older (responders: 12, non-responders: 35), younger (responders: 9, non-responders: 42), male (responders: 12, non-responders: 45), and female (responders: 9, non-responders: 32) participants). The dataset was randomly divided into a training set (80% of samples) and a testing set (20% of samples). Within the training set, we first assessed class balance. Given the imbalance of sample size in responder and non-responder classes, we applied the SMOTE algorithm (DMwR, v0.4.1)^95^ to balance the samples. Subsequently, 5-fold cross-validation was performed to identify the optimal model. The best model was then evaluated on the training set to compute overall model performance. To enhance the reliability of the evaluation, this resampling and training procedure was repeated 100 times. Model performance was assessed using the area under the ROC curve (AUC), and the final AUC reported represents the mean of the 100 iterations. Feature importance was evaluated using the Mean Decrease Gini, with the reported importance values similarly representing the average value across all 100 iterations.

The above procedure was performed in all participants at the baseline and in each subgroup (by age: >58 vs. ≤58; by gender: male vs. female) using single or multi-omics datasets as the input (clinical, microbiome, metabolite, clinical + microbiome, clinical + metabolite, and clinical + microbiome + metabolite).

## Declarations

### Ethics approval and consent to participate

This study was conducted with approval from the IRB of UConn Health in accordance with the relevant guidelines and regulations. Participants were recruited from the community and consented to participation in a double-blind, randomized, placebo-controlled study investigating the use of dutasteride in reducing drinking (ClinicalTrials.gov identifier: NCT04098302, Dutasteride Treatment for Reducing Heavy Drinking in AUD: Predictors of Efficacy, first registered 19/09/2019). The study protocol is published online.^1^

### Consent for publication

Not applicable. The manuscript does not contain data from any individual person.

### Availability of data and material

Raw sequencing reads from 16S rRNA gene amplicon sequencing are available *via* the National Center for Biotechnology Information (NCBI) Short Read Archive (SRA) under accession number PRJNA1401844.

## Authors’ contributions

JMC, HT, and YZ were responsible for study design. LRD, DJL, QL, HY, JMC, and HG performed data analysis. LRD, DJL, and YZ interpreted findings and drafted the manuscript. All authors critically reviewed the manuscript and agree to the publication of the final version.

## Data Availability

Raw sequencing reads from 16S rRNA gene amplicon sequencing are available via the National Center for Biotechnology Information (NCBI) Short Read Archive (SRA) under accession number PRJNA1401844.

## Acknowledgements

This work was supported by the National Institute on Alcohol Abuse and Alcoholism (NIAAA) under grants T32 AA007290 and P50 AA027055 and by the National Institute of Health (NIH) General Clinical Research Centers Program under grant M01 RR0619. We thank Judy Kalinowski at the UConn Health Clinical Research Center for stool and serum sample management.

## Ethics approval and consent to participate

This study was conducted with approval from the UConn Health IRB in accordance with the relevant guidelines and regulations. Participants were recruited from the community and consented to participation in a double-blind, randomized, placebo-controlled study investigating the use of dutasteride in reducing drinking (ClinicalTrials.gov identifier: NCT04098302, Dutasteride Treatment for Reducing Heavy Drinking in AUD: Predictors of Efficacy, first registered 19/09/2019). The study protocol is published online.^1^

## Consent for publication

Not applicable. The manuscript does not contain data from any individual person.

---

**Supporting Figure 1:**
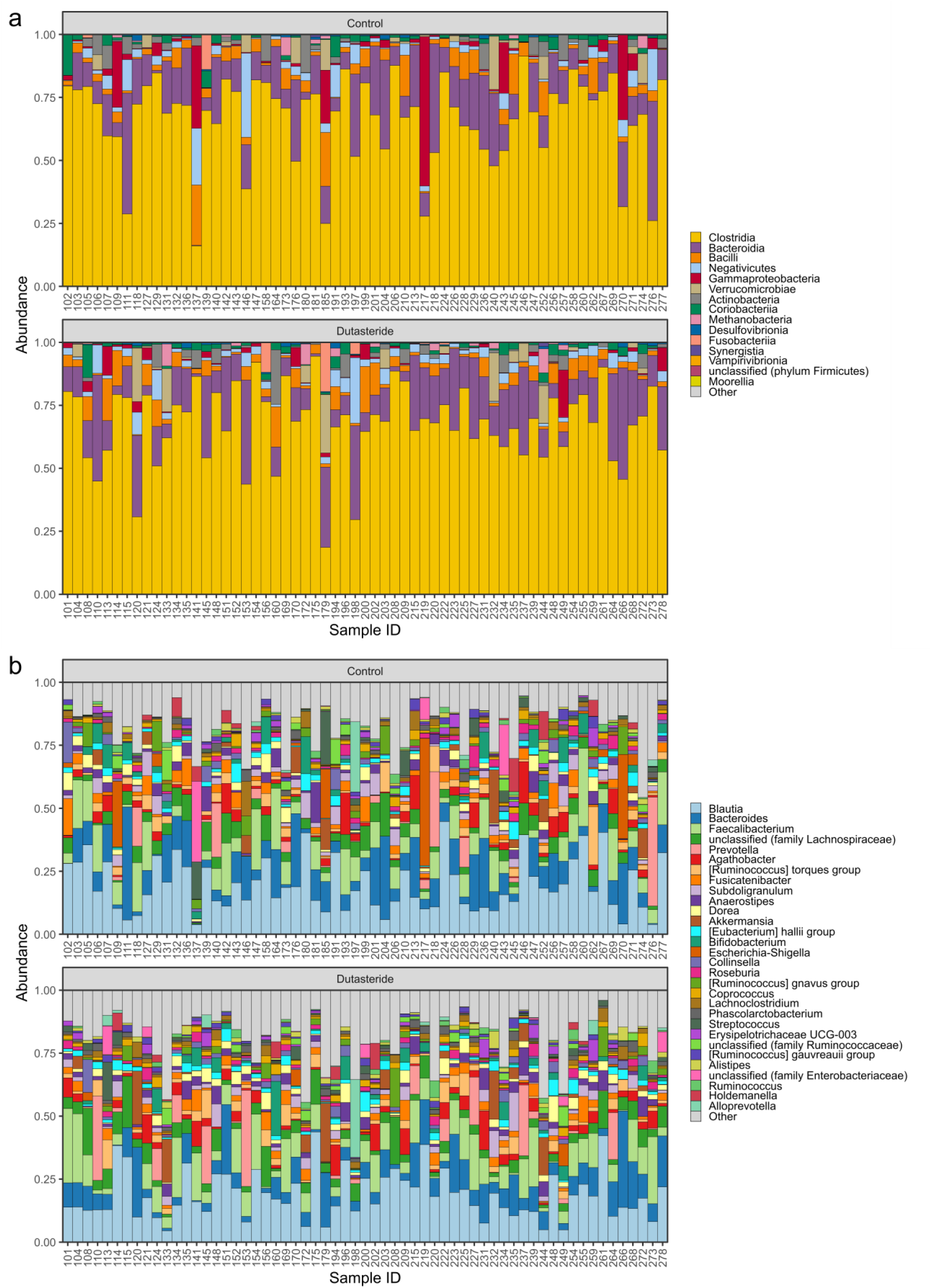
Relative abundance bar plots of the baseline samples separated by treatment group. Microbiome feature abundances are shown for each sample ID at class (a) and genus (b) level. Samples in the control and dutasteride treatment group are separated by the plot facets.

**Supporting Figure 2:**
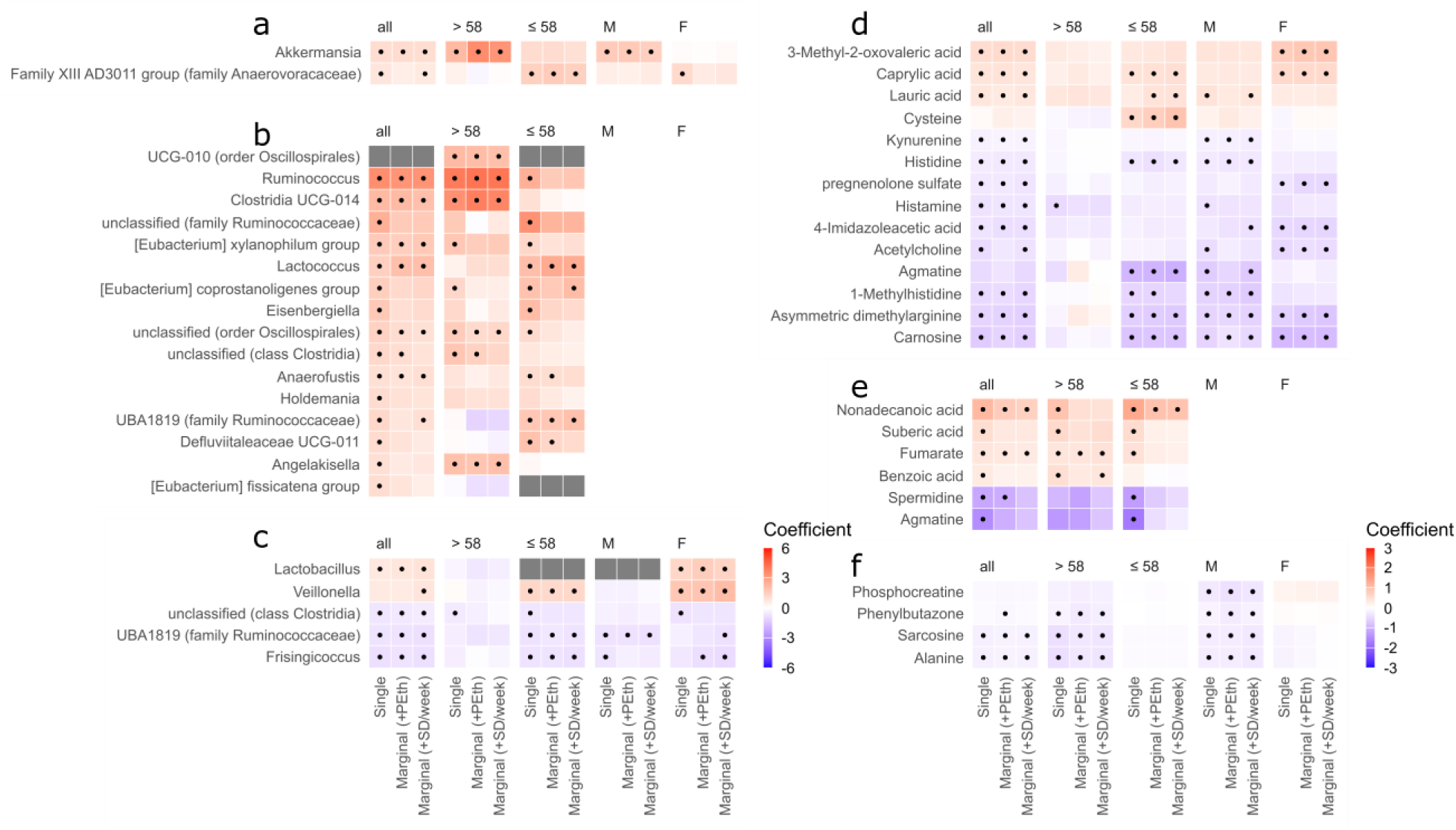
Microbiome and metabolite changes associated with age, sex, and BMI. (a-c) Heatmap of associations with individual microbiome features. Age is shown in (a), sex is shown in (b), and BMI is shown in (c). (d-f) Heatmap of associations with individual metabolite features. Age is shown in (d), sex is shown in (e), and BMI is shown in (f). In all plots, “single” on the x-axis refers to significance in a single-variable model (e.g. feature vs. age with no covariates), and “marginal” refers to the effect of that variable controlling for covariates, using the same GLM models in Figures 2 and 3. Subgroups are shown in plot facets, and dots indicate statistically significant associations (q < 0.1). Note that the sex subgroups are absent from (b) and (e) because all participants in the male and female subgroups share an identical value for sex. The age subgroups are still present in (a) and (d) because participants have unique values for age that enable comparison.

**Supporting Figure 3:**
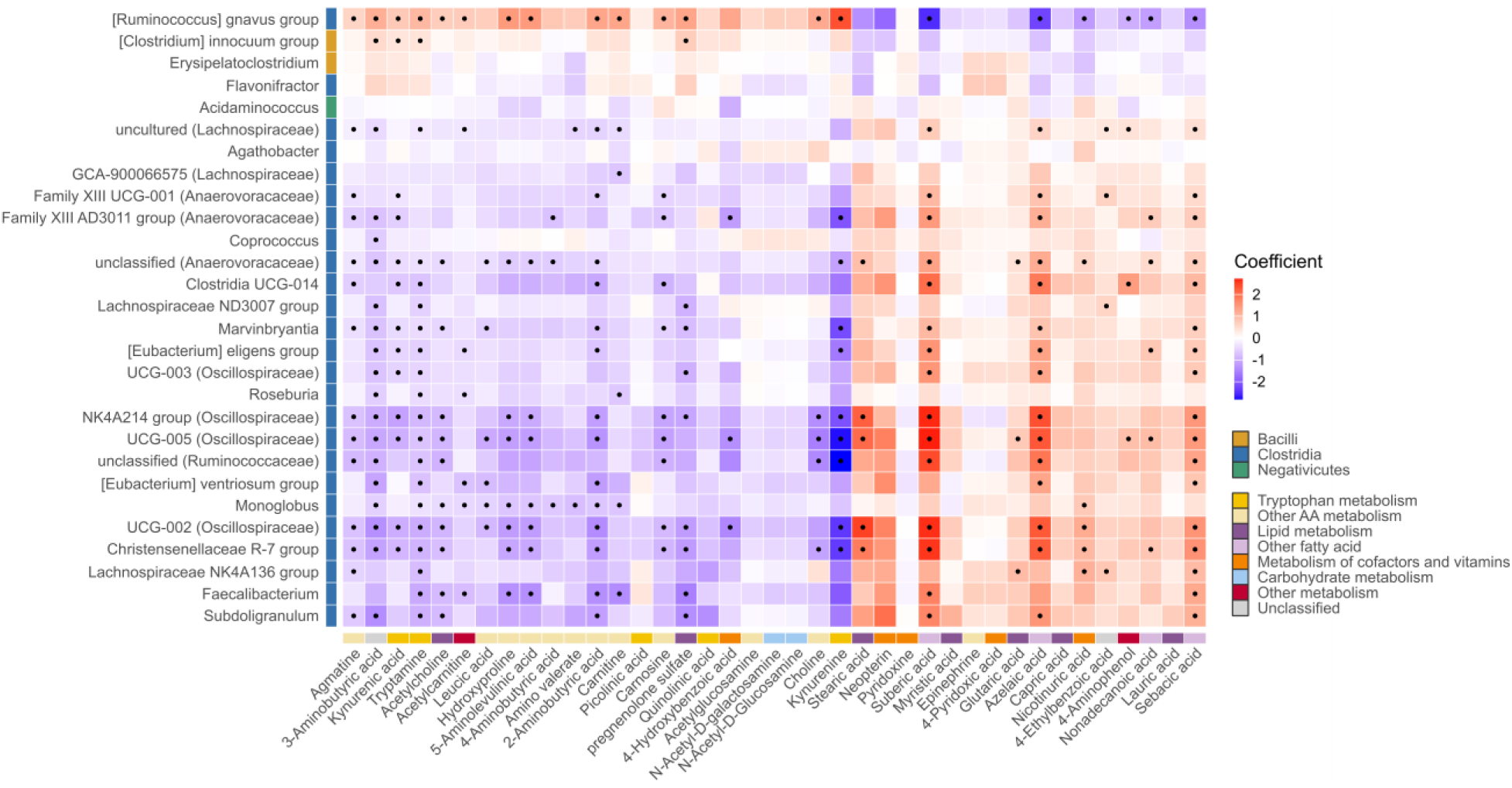
Heatmap of microbiome-metabolome co-associations. Heatmap showing direct correlations between metabolite and microbiome features. Values and methods are identical to the chord diagram shown in Figure 4a and 4b. Color keys on the left indicate the class of the microbiome feature, and those on the bottom indicate the KEGG pathway classification of the metabolite. Dots indicate statistically significant associations (q < 0.1).

**Supporting Figure 4:**
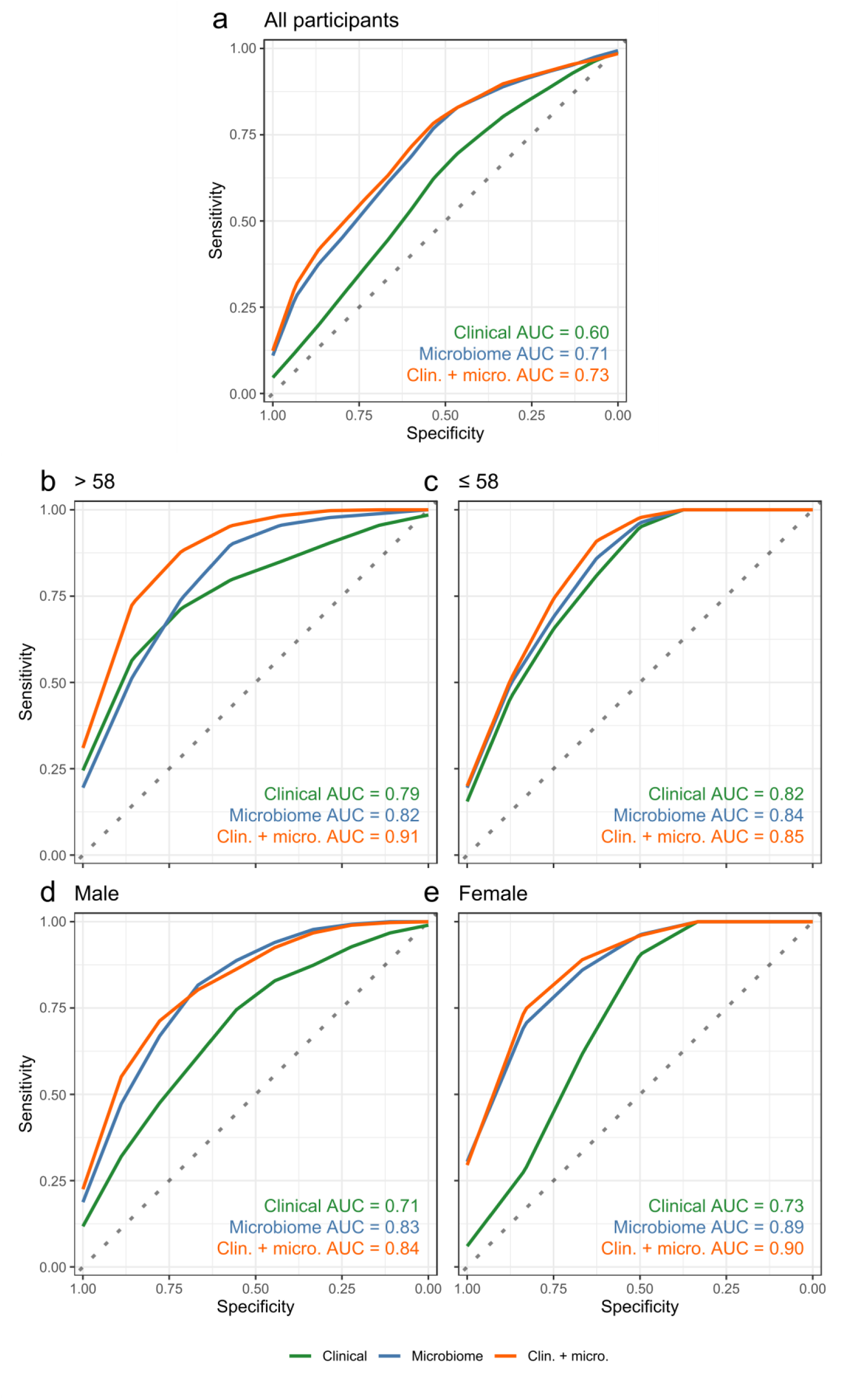
Microbiome-based random forest classifier performance for drinking response. Comparing a clinical + microbiome classifier to classifiers using each dataset alone. Refer to Figure 5 for plot information.

**Supporting Figure 5:**
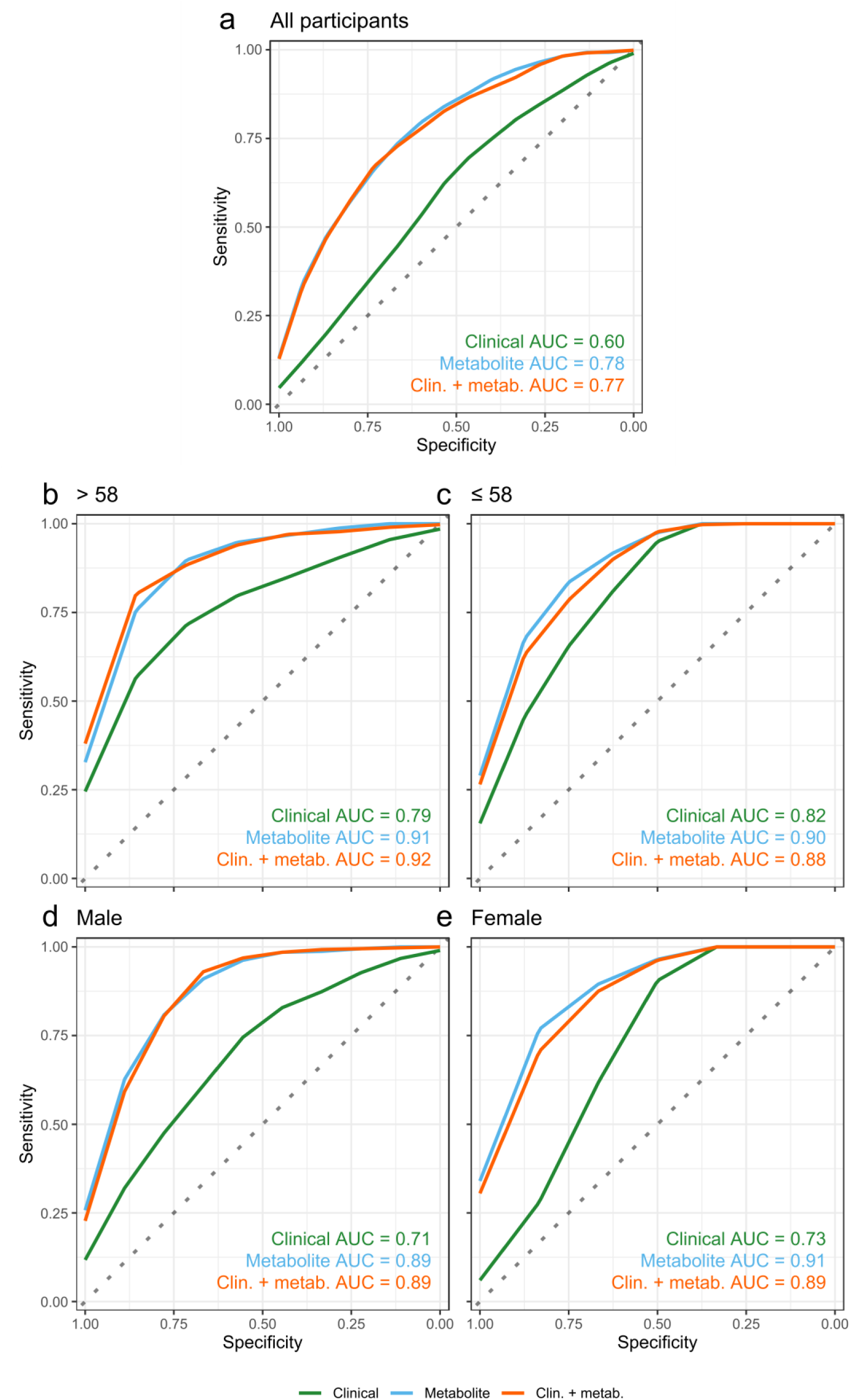
Metabolite-based random forest classifier performance for drinking response. Comparing a clinical + metabolite classifier to classifiers using each dataset alone. Refer to Figure 5 for plot information.

**Supporting Figure 6:**
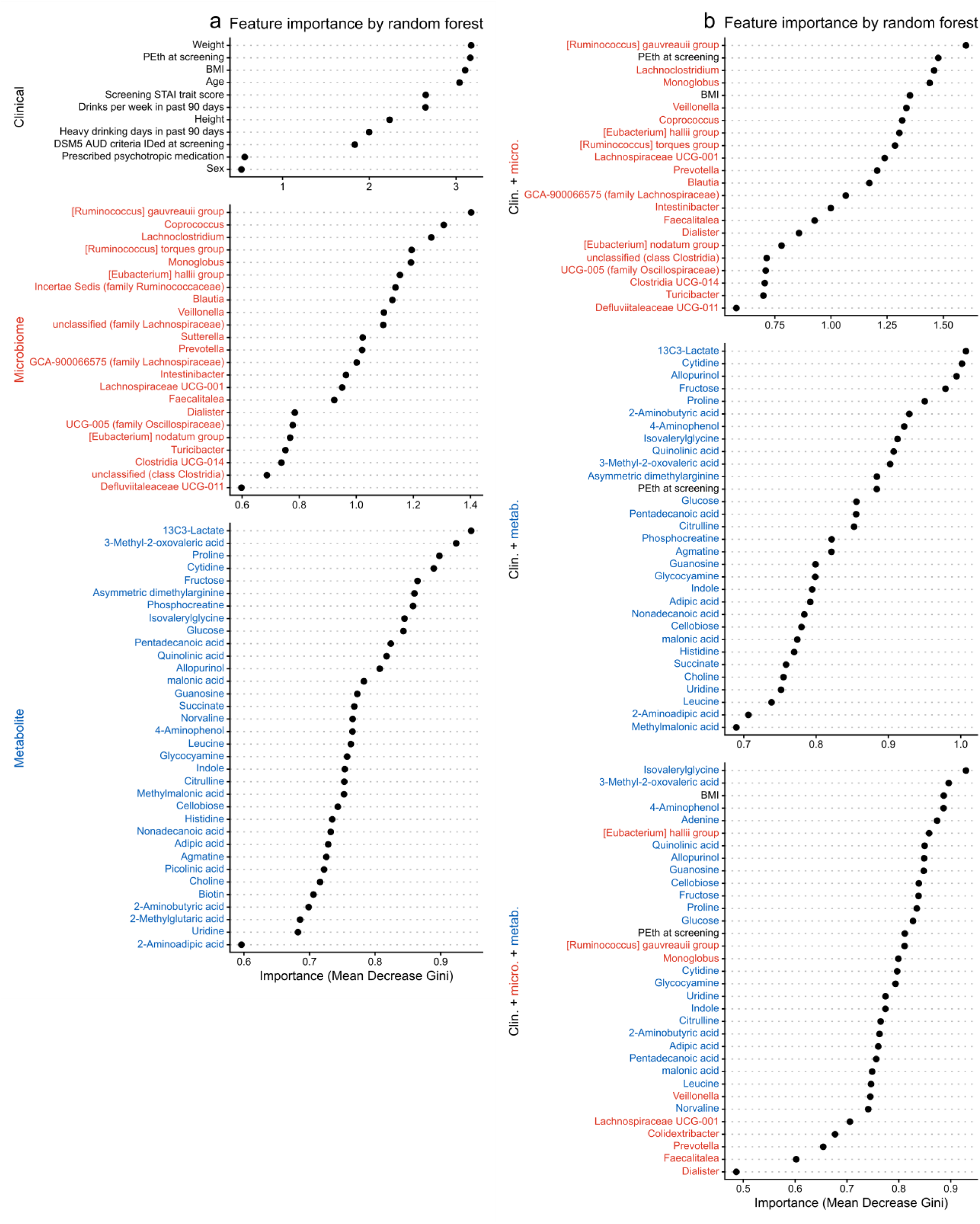
RF classifier feature importance in the full cohort. X-axis values indicate the Mean Decrease Gini for each predictor variable included the model. Higher values indicate that a given variable is more relevant to separation by class. Feature importance is shown for the models including one dataset (a) and multiple datasets (b). Feature names are colored by their source dataset: black for clinical, red for microbiome, and blue for metabolites.

**Supporting Figure 7:**
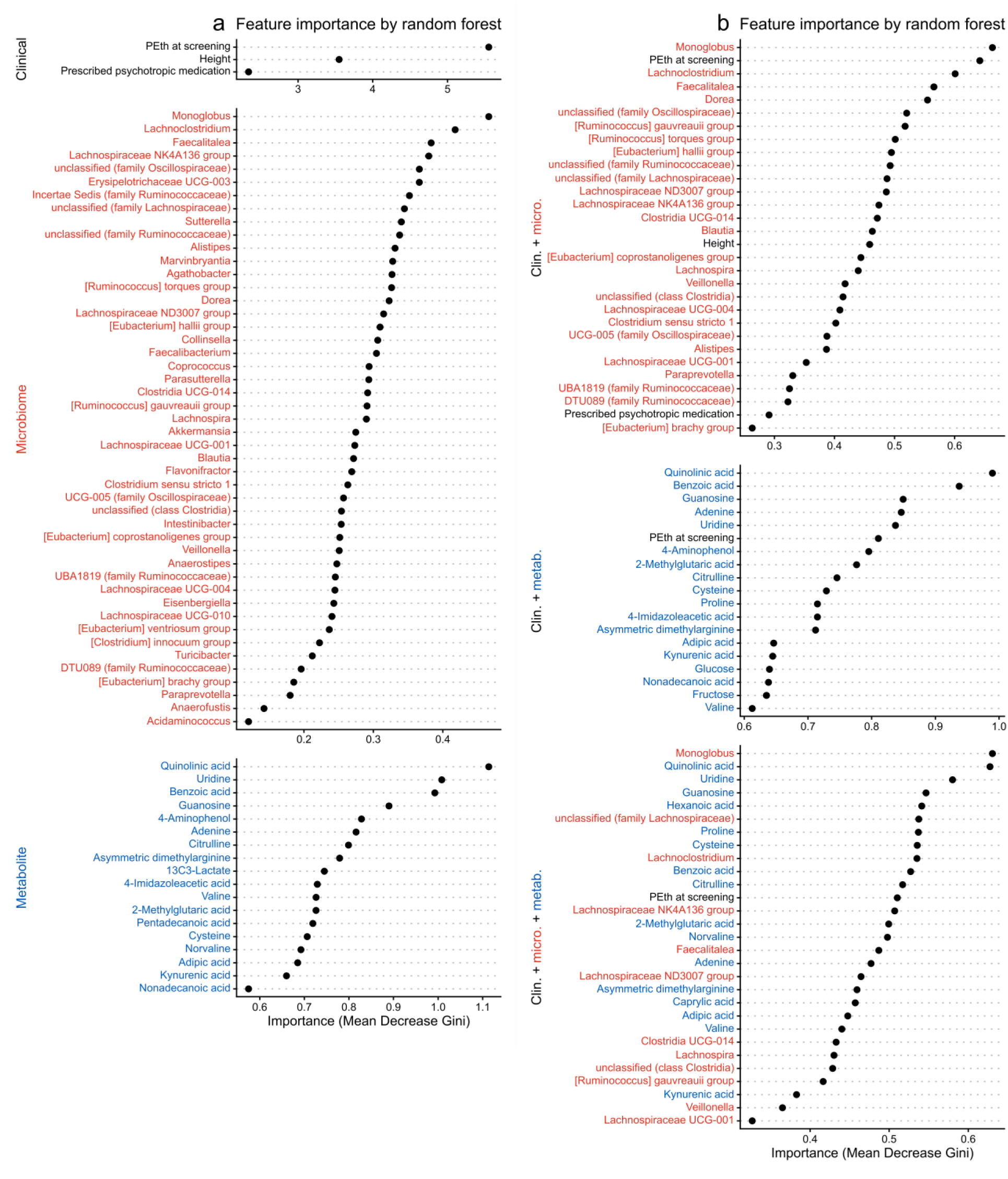
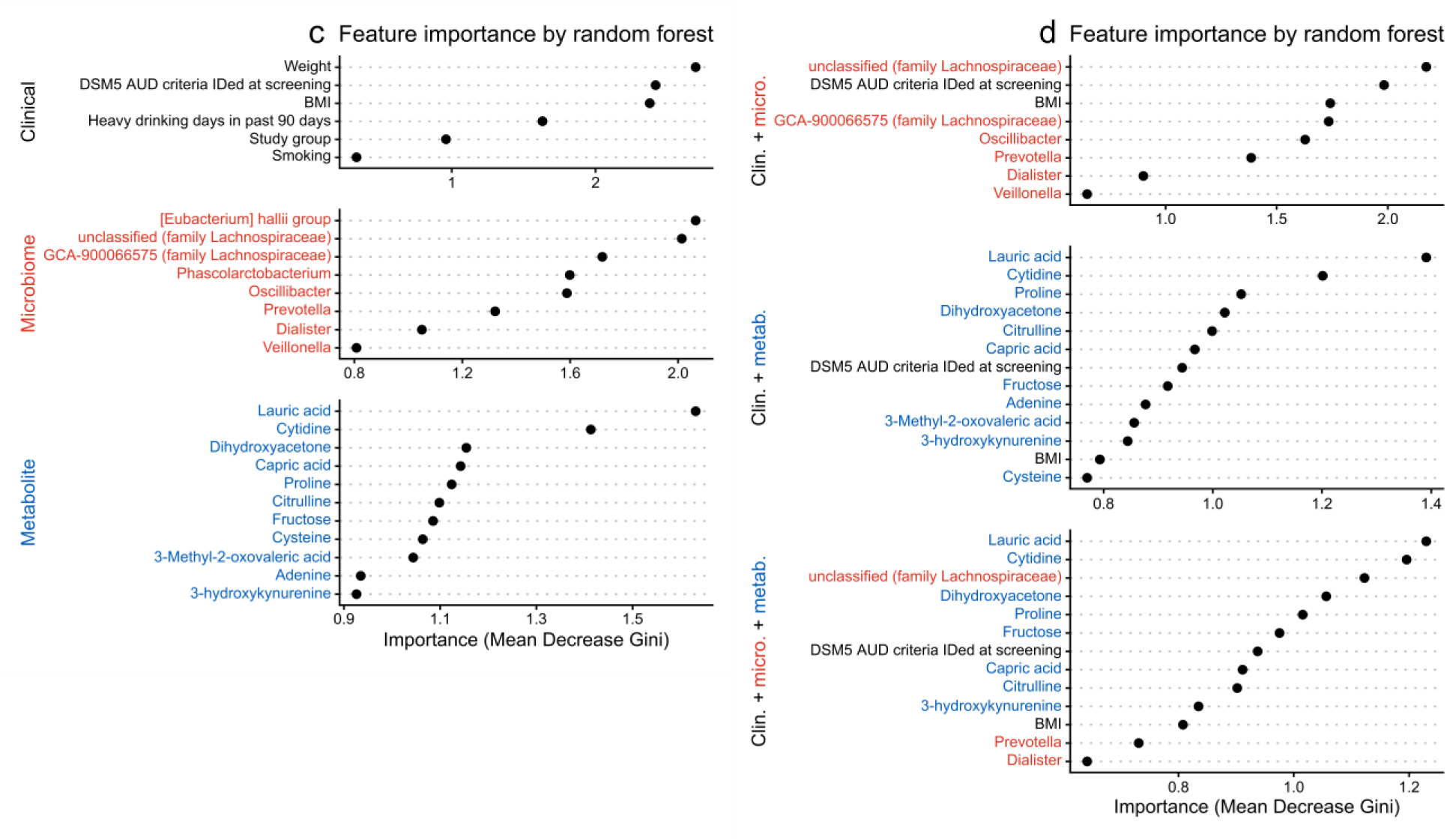
RF classifier feature importance in age subgroups. Feature importance plots show results for single (a) and multiple (b) datasets in the participants over 58 years old, and single (a) and multiple (b) datasets in the participants aged 58 years and younger. Refer to Supporting Figure 4 for plot information.

**Supporting Figure 8:**
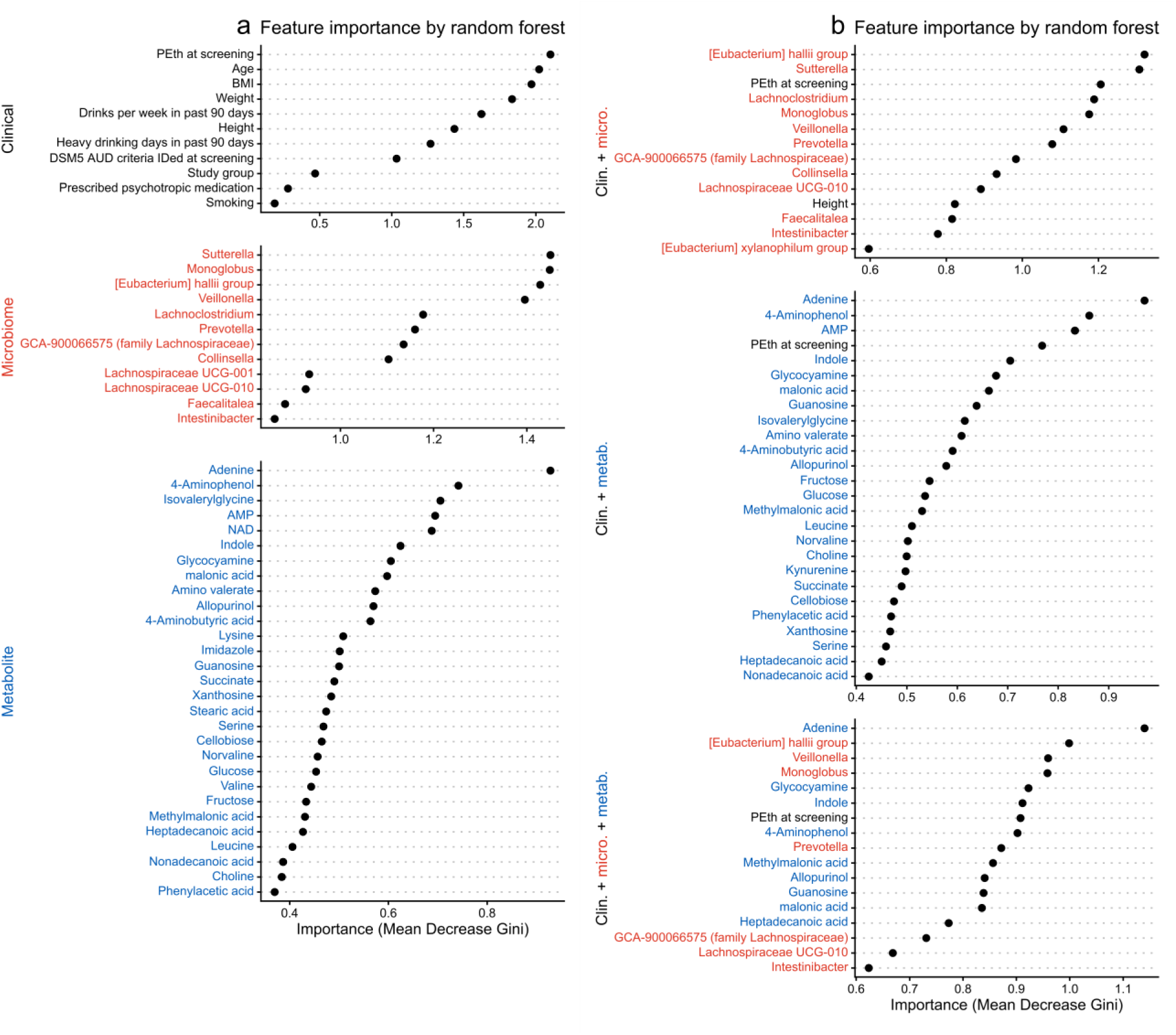
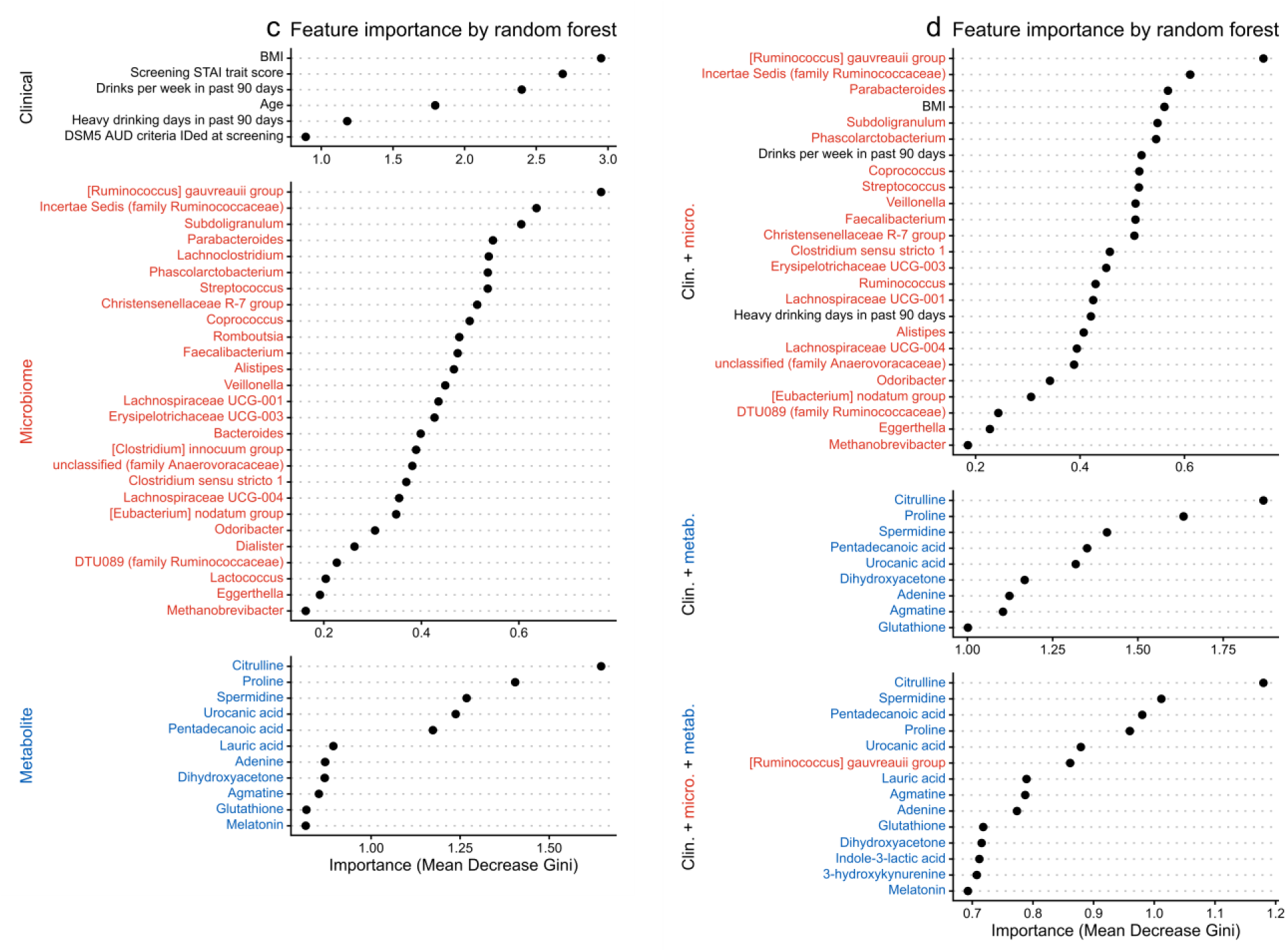
RF classifier feature importance in sex subgroups. Feature importance plots show results for single (a) and multiple (b) datasets in the male participants, and single (a) and multiple (b) datasets in the female participants. Refer to Supporting Figure 4 for plot information.

**Supporting Table 1:** Demographic and clinical factors for young and old subgroups.

| Variable | Overall<br>N = 122<sup>1</sup> | >58 Years<br>N = 60<sup>1</sup> | ≤58 Years<br>N = 62<sup>1</sup> | p-value |
| --- | --- | --- | --- | --- |
| **Treatment group, n, (%)** |  |  |  | 0.4<sup>2</sup> |
| Placebo | 61, (50%) | 27, (45%) | 34, (55%) |  |
| Dutasteride | 61, (50%) | 26, (57%) | 35, (46%) |  |
| **Sex, n, (%)** |  |  |  | 0.2<sup>2</sup> |
| Male | 76, (62%) | 41, (68%) | 35, (56%) |  |
| Female | 46, (38%) | 19, (32%) | 27, (44%) |  |
| **Age, Mean, (SD)** | 57, (9) | 65, (3) | 49, (6) | <0.001<sup>3</sup> |
| **BMI, Mean, (SD)** | 29.3, (5.8) | 28.6, (5.1) | 30.0, (6.4) | 0.2<sup>3</sup> |
| **Prescribed psychotropic medication, n, (%)** | 42, (34%) | 21, (35%) | 21, (34%) | >0.9<sup>2</sup> |
| **Smoking, n, (%)** | 15, (12%) | 6, (10%) | 9, (15%) | 0.6<sup>2</sup> |
| **Primary race, n, (%)** |  |  |  | 0.7<sup>2</sup> |
| White | 115, (94%) | 56, (93%) | 59, (95%) |  |
| Black | 7, (5.7%) | 4, (6.7%) | 3, (4.8%) |  |
| **Screening STAI trait Score, Mean, (SD)** | 39, (12) | 36, (11) | 41, (12) | 0.017<sup>3</sup> |
| **DSM5 AUD criteria identified at screening, Mean, (SD)** | 5.75, (2.29) | 5.00, (2.06) | 6.47, (2.29) | <0.001<sup>3</sup> |
| **PEth at screening, Mean, (SD)** | 353, (290) | 312, (262) | 392, (312) | 0.14<sup>3</sup> |
| **Average drinks per week at screening, Mean, (SD)** | 45, (22) | 43, (18) | 46, (25) | 0.4<sup>3</sup> |

<sup>1</sup>Frequency, (%) or Mean, (SD)
<sup>2</sup>Fisher's exact test
<sup>3</sup>Welch Two Sample t-test

**Supporting Table 2:** Demographic and clinical factors for male and female subgroups.

| Variable | Overall<br>N = 122<sup>1</sup> | Male<br>N = 76<sup>1</sup> | Female<br>N = 46<sup>1</sup> | p-value |
| --- | --- | --- | --- | --- |
| **Treatment group, n, (%)** |  |  |  | 0.4<sup>2</sup> |
| Placebo | 61, (50%) | 41, (54%) | 20, (43%) |  |
| Dutasteride | 61, (50%) | 35, (46%) | 26, (57%) |  |
| **Age, Mean, (SD)** | 57, (9) | 57, (9) | 57, (9) | 0.8<sup>3</sup> |
| **BMI, Mean, (SD)** | 29.3, (5.8) | 29.9, (4.8) | 28.4, (7.2) | 0.2<sup>3</sup> |
| **Prescribed psychotropic medication, n, (%)** | 42, (34%) | 19, (25%) | 23, (50%) | 0.006<sup>2</sup> |
| **Smoking, n, (%)** | 15, (12%) | 9, (12%) | 6, (13%) | >0.9<sup>2</sup> |
| **Primary race, n, (%)** |  |  |  | 0.4<sup>2</sup> |
| White | 115, (94%) | 73, (96%) | 42, (91%) |  |
| Black | 7, (5.7%) | 3, (3.9%) | 4, (8.7%) |  |
| **Screening STAI trait score, Mean, (SD)** | 39, (12) | 38, (11) | 40, (13) | 0.2<sup>3</sup> |
| **DSM5 AUD criteria identified at screening, Mean, (SD)** | 5.75, (2.29) | 5.83, (2.42) | 5.61, (2.08) | 0.6<sup>3</sup> |
| **PEth at screening, Mean, (SD)** | 353, (290) | 371, (299) | 325, (278) | 0.4<sup>3</sup> |
| **Average drinks per week at screening, Mean, (SD)** | 45, (22) | 51, (23) | 35, (15) | <0.001<sup>3</sup> |

<sup>1</sup>Frequency, (%) or Mean, (SD)
<sup>2</sup>Fisher's exact test
<sup>3</sup>Welch Two Sample t-test

**Supporting Table 3:** Demographic and clinical factors for non-responders and responders with endpoint clinical data.

| Variable | Overall<br>N = 98<sup>1</sup> | Non-responder<br>N = 77<sup>1</sup> | Responder<br>N = 21<sup>1</sup> | p-value |
| --- | --- | --- | --- | --- |
| **Treatment group, n, (%)** |  |  |  | 0.15<sup>2</sup> |
| Placebo | 47, (48%) | 40, (52%) | 7, (33%) |  |
| Dutasteride | 51, (52%) | 37, (48%) | 14, (67%) |  |
| **Sex, n, (%)** |  |  |  | >0.9<sup>2</sup> |
| Male | 57, (58%) | 45, (58%) | 12, (57%) |  |
| Female | 41, (42%) | 32, (42%) | 9, (43%) |  |
| **Age, Mean, (SD)** | 57, (9) | 56, (9) | 58, (11) | 0.6<sup>3</sup> |
| **BMI, Mean, (SD)** | 28.9, (5.8) | 28.4, (5.0) | 30.3, (8.3) | 0.3<sup>3</sup> |
| **Prescribed psychotropic medication, n, (%)** | 35, (36%) | 29, (38%) | 6, (29%) | 0.6<sup>2</sup> |
| **Smoking, n, (%)** | 13, (13%) | 11, (14%) | 2, (9.5%) | 0.7<sup>2</sup> |
| **Primary race, n, (%)** |  |  |  | 0.6<sup>2</sup> |
| White | 92, (94%) | 73, (95%) | 19, (90%) |  |
| Black | 6, (6.1%) | 4, (5.2%) | 2, (9.5%) |  |
| **Screening STAI trait score, Mean, (SD)** | 39, (11) | 39, (11) | 39, (14) | >0.9<sup>3</sup> |
| **DSM5 AUD criteria identified at screening, Mean, (SD)** | 5.69, (2.27) | 5.83, (2.26) | 5.19, (2.32) | 0.3<sup>3</sup> |
| **PEth at screening, Mean, (SD)** | 367, (303) | 387, (307) | 294, (282) | 0.2<sup>3</sup> |
| **PEth at endpoint, Mean, (SD)** | 296, (281) | 344, (292) | 125, (146) | <0.001<sup>3</sup> |
| **Average drinks per week at screening, Mean, (SD)** | 44, (22) | 44, (23) | 44, (17) | >0.9<sup>3</sup> |
| **Average drinks per week at endpoint, Mean, (SD)** | 30, (21) | 36, (19) | 7, (7) | <0.001<sup>3</sup> |
| **Percent reduction in PEth, Mean, (SD)** | 18, (51) | 8, (42) | 51, (67) | 0.010<sup>3</sup> |
| **Percent drinking reduction, Mean, (SD)** | 29, (38) | 15, (28) | 84, (16) | <0.001<sup>3</sup> |

<sup>1</sup>Frequency (%) or Mean (SD)
<sup>2</sup>Fisher's exact test
<sup>3</sup>Welch Two Sample t-test

**Supporting Table 4:** Demographic and clinical factors for baseline-only and paired samples.

| Variable | Overall<br>N = 122<sup>1</sup> | Baseline Only<br>N = 45<sup>1</sup> | Baseline &<br>Endpoint<br>N = 77<sup>1</sup> | p-value |
| --- | --- | --- | --- | --- |
| **Treatment group, n, (%)** |  |  |  | 0.7<sup>2</sup> |
| Placebo | 61, (50%) | 24, (53%) | 37, (48%) |  |
| Dutasteride | 61, (50%) | 21, (47%) | 40, (52%) |  |
| **Sex, n, (%)** |  |  |  | 0.3<sup>2</sup> |
| Male | 76, (62%) | 31, (69%) | 45, (58%) |  |
| Female | 46, (38%) | 14, (31%) | 32, (42%) |  |
| **Age, Mean, (SD)** | 57, (9) | 56, (9) | 57, (9) | 0.5<sup>3</sup> |
| **BMI, Mean, (SD)** | 29.3, (5.8) | 30.7, (5.8) | 28.5, (5.7) | 0.041<sup>3</sup> |
| **Prescribed psychotropic medication, n, (%)** | 42, (34%) | 14, (31%) | 28, (36%) | 0.7<sup>2</sup> |
| **Smoking, n, (%)** | 15, (12%) | 5, (11%) | 10, (13%) | >0.9<sup>2</sup> |
| **Primary race, n, (%)** |  |  |  | 0.10<sup>2</sup> |
| White | 115, (94%) | 40, (89%) | 75, (97%) |  |
| Black | 7, (5.7%) | 5, (11%) | 2, (2.6%) |  |
| **Screening STAI trait score, Mean, (SD)** | 39, (12) | 41, (13) | 38, (11) | 0.2<sup>3</sup> |
| **DSM5 AUD criteria identified at screening, Mean, (SD)** | 6, (2) | 6, (3) | 5, (2) | 0.086<sup>3</sup> |
| **PEth at screening, Mean, (SD)** | 353, (290) | 372, (317) | 343, (277) | 0.6<sup>3</sup> |
| **PEth at endpoint, Mean, (SD)** | 301, (284) | 364, (365) | 282, (254) | 0.3<sup>3</sup> |
| **Average drinks per week at screening, Mean, (SD)** | 45, (22) | 45, (19) | 45, (23) | >0.9<sup>3</sup> |
| **Average drinks per week at endpoint, Mean, (SD)** | 30, (21) | 33, (26) | 29, (18) | 0.6<sup>3</sup> |
| **Percent reduction in PEth, Mean, (SD)** | 18, (51) | 27, (44) | 14, (53) | 0.2<sup>3</sup> |
| **Percent reduction in drinks per week, Mean, (SD)** | 33, (31) | 36, (34) | 32, (30) | 0.6<sup>3</sup> |

<sup>1</sup>Frequency, (%) or Mean, (SD)
<sup>2</sup>Fisher's exact test
<sup>3</sup>Welch Two Sample t-test

**Supporting Table 5:** Demographic and clinical factors for paired placebo and treatment samples.

| Variable | Overall<br>N = 77<sup>1</sup> | Placebo<br>N = 37<sup>1</sup> | Dutasteride<br>N = 40<sup>1</sup> | p-value |
| --- | --- | --- | --- | --- |
| **Sex, n, (%)** |  |  |  | >0.9<sup>2</sup> |
| Male | 45, (58%) | 22, (59%) | 23, (58%) |  |
| Female | 32, (42%) | 15, (41%) | 17, (43%) |  |
| **Age, Mean, (SD)** | 57, (9) | 57, (9) | 57, (10) | >0.9<sup>3</sup> |
| **BMI, Mean, (SD)** | 28.5, (5.7) | 27.9, (4.5) | 29.0, (6.6) | 0.4<sup>3</sup> |
| **Prescribed psychotropic medication, n, (%)** | 28, (36%) | 8, (22%) | 20, (50%) | 0.017<sup>2</sup> |
| **Smoking, n, (%)** | 10, (13%) | 5, (14%) | 5, (13%) | >0.9<sup>2</sup> |
| **Primary race, n, (%)** |  |  |  | 0.5<sup>2</sup> |
| White | 2, (2.6%) | 0, (0%) | 2, (5.0%) |  |
| Black | 75, (97%) | 37, (100%) | 38, (95%) |  |
| **Screening STAI trait score, Mean, (SD)** | 38, (11) | 36, (10) | 39, (11) | 0.14<sup>3</sup> |
| **DSM5 AUD criteria identified at screening, Mean, (SD)** | 5, (2) | 5, (2) | 5, (2) | 0.9<sup>3</sup> |
| **PEth at screening, Mean, (SD)** | 343, (277) | 312, (280) | 370, (275) | 0.4<sup>3</sup> |
| **PEth at endpoint, Mean, (SD)** | 282, (254) | 278, (258) | 285, (253) | 0.9<sup>3</sup> |
| **Average drinks per week at screening, Mean, (SD)** | 45, (23) | 45, (25) | 45, (23) | >0.9<sup>3</sup> |
| **Average drinks per week at endpoint, Mean, (SD)** | 29, (18) | 29, (19) | 29, (18) | >0.9<sup>3</sup> |
| **Percent reduction in PEth, Mean, (SD)** | 14, (53) | 9, (56) | 19, (50) | 0.4<sup>3</sup> |
| **Percent reduction in drinks per week, Mean, (SD)** | 32, (30) | 34, (30) | 31, (30) | 0.7<sup>3</sup> |

<sup>1</sup>Frequency, (%) or Mean, (SD)
<sup>2</sup>Fisher's exact test
<sup>3</sup>Welch Two Sample t-test

**Supporting Table 6:** Demographic and clinical factors for non-responders and responders with endpoint microbiome data.

| Variable | Overall<br>N = 77<sup>1</sup> | Non-responder<br>N = 62<sup>1</sup> | Responder<br>N = 15<sup>1</sup> | p-value |
| --- | --- | --- | --- | --- |
| **Treatment group, n, (%)** |  |  |  | >0.9<sup>2</sup> |
| Placebo | 37, (48%) | 30, (48%) | 7, (47%) |  |
| Dutasteride | 40, (52%) | 32, (52%) | 8, (53%) |  |
| **Sex, n, (%)** |  |  |  | >0.9<sup>2</sup> |
| Male | 45, (58%) | 36, (58%) | 9, (60%) |  |
| Female | 32, (42%) | 26, (42%) | 6, (40%) |  |
| **Age, Mean, (SD)** | 57, (9) | 57, (9) | 58, (11) | 0.9<sup>3</sup> |
| **BMI, Mean, (SD)** | 28.5, (5.7) | 27.9, (5.1) | 30.8, (7.7) | 0.2<sup>3</sup> |
| **Prescribed psychotropic medication, n, (%)** | 28, (36%) | 25, (40%) | 3, (20%) | 0.2<sup>2</sup> |
| **Smoking, n, (%)** | 10, (13%) | 9, (15%) | 1, (6.7%) | 0.7<sup>2</sup> |
| **Primary race, n, (%)** |  |  |  | 0.4<sup>2</sup> |
| White | 75, (97%) | 61, (98%) | 14, (93%) |  |
| Black | 2, (2.6%) | 1, (1.6%) | 1, (6.7%) |  |
| **Screening STAI trait score, Mean, (SD)** | 38, (11) | 39, (11) | 33, (8) | 0.027<sup>3</sup> |
| **DSM5 AUD criteria identified at screening, Mean, (SD)** | 5.45, (2.04) | 5.65, (2.05) | 4.67, (1.88) | 0.089<sup>3</sup> |
| **PEth at screening, Mean, (SD)** | 343, (277) | 354, (271) | 297, (307) | 0.5<sup>3</sup> |
| **PEth at endpoint, Mean, (SD)** | 282, (254) | 320, (260) | 129, (152) | <0.001<sup>3</sup> |
| **Average drinks per week at screening, Mean, (SD)** | 45, (23) | 44, (25) | 47, (19) | 0.7<sup>3</sup> |
| **Average drinks per week at endpoint, Mean, (SD)** | 29, (18) | 35, (16) | 8, (7) | <0.001<sup>3</sup> |
| **Percent reduction in PEth, Mean, (SD)** | 14, (53) | 6, (43) | 46, (74) | 0.061<sup>3</sup> |
| **Percent reduction in drinks per week, Mean, (SD)** | 30, (33) | 18, (22) | 81, (16) | <0.001<sup>3</sup> |

<sup>1</sup>Frequency (%) or Mean (SD)
<sup>2</sup>Fisher's exact test
<sup>3</sup>Welch Two Sample t-test

